# Stool host transcriptome reveals gut epithelial cell states and immune dynamics for gastrointestinal diagnosis and monitoring

**DOI:** 10.64898/2026.10.31.26366938

**Authors:** Wenying Pan, Xiaxi Li, Ziyu Yang, Dan Chen, Yanyang Liang, Haijiu Lin, Jing Huang, Shuyu He, Boxuan Liu, Changpu Song, Min Li

**Affiliations:** Guangdong Jiyin Biotech, D3 Building, TCL international E city, no. 1001, Zhong Shan Yuan Road, Nanshan District. Shenzhen, China; Department of Gastroenterology, Shenzhen Hospital, Southern Medical University, Shenzhen, China; Department of Laboratory Medicine, Renji Hospital, School of Medicine, Shanghai Jiao Tong University, Shanghai, China

## Abstract

Gastrointestinal diagnosis and monitoring rely on colonoscopy, which requires anesthesia and a long wait, and non-invasive alternatives are lacking. Here we show that whole-transcriptome sequencing of stool host RNA, recovering a median of 6,300 protein-coding genes across 285 libraries spanning colorectal cancer (CRC) and inflammatory bowel disease (IBD), precancerous lesions (APL), and normal samples, reads a census of the cells shed into the lumen. In normal samples, epithelium supplies 81% and immune cells supply 11% of stool host mRNA transcripts; the immune cells’ share rises in CRC and IBD, and B- and T-cell receptor clonotypes rise with it. In this work, we show that CRC, IBD, and normal samples carry distinct transcriptome profiles, which a three-class model separates with an area under the curve of 0.94–0.96, and that within inflammatory bowel disease a 15-gene neutrophil score tracks endoscopic activity better than fecal calprotectin in the same stool.

## Introduction

Gastrointestinal diagnosis and monitoring rely heavily on colonoscopy, which requires bowel preparation and anesthesia, and carries a small procedural risk. A patient presenting with non-red-flag gastrointestinal symptoms such as abdominal pain, diarrhea or bloating is worked up by exclusion—blood counts, C-reactive protein, stool culture, fecal calprotectin, celiac serology— before colonoscopy settles whether the cause is neoplastic, inflammatory or functional ^1,2^. The diagnostic pathway can take weeks to months. Once inflammatory bowel disease (IBD) is diagnosed, activity is monitored by repeat endoscopy or by fecal calprotectin, a single neutrophil protein that reports inflammation as one number and correlates only moderately with mucosal healing, particularly in Crohn’s disease ^3,4^. These two clinical problems share one missing measurement: a molecular readout of the state of the gut tissue and its immune response, obtainable repeatedly from a sample the patient can collect at home.

Stool contains that readout. Epithelial cells exfoliate continuously along the entire tract, and immune cells that traffic into the mucosa are shed with them, so the human mRNA in stool integrates active biology across the intestine rather than reporting a single biopsy site or the systemic average that blood provides ^5,6^. Recovering human RNA in stool is hard: most stool RNA is microbial, RNase activity is high, PCR inhibitors are abundant, and the matrix is hard to homogenize reproducibly ^7,8^. The field has taken two routes around the microbial background. The first is to sequence only chosen genes. Huang et al. used amplicon sequencing (Foli-seq) of 543 genes and characterized immune dynamics in the healthy and inflamed gut ^9^. Ghannam et al. used hybridization capture of a custom 1,507-transcript panel from stool RNA to separate active from quiescent Crohn’s disease ^10^. These assays read the genes they were built to read: they cannot report a compartment that was not targeted, cannot assign transcripts to cell types across a whole reference atlas, and cannot be asked a new question without a new design. The second route is poly(A) capture. Using this approach, Halpern et al. recovered a median of 1,745 human genes per stool sample at ∼20 million reads, and distinguished CRC patients from controls with a signal that mirrored the matched tumor tissue ^11^. These studies show the diagnostic potential of host mRNA in stool, but the genes they detect are a small fraction of the roughly twenty thousand protein-coding genes in the human genome, and their cohorts were either one disease at varying activity or one disease against healthy controls. Whether the stool host transcriptome can tell two gastrointestinal diseases apart has not been explored. Likewise, previous stool-mRNA studies have separated active IBD from remission using a classification model, but none has attempted to track the dynamic changes of different types of immune cells and immune repertoire as the disease severity of IBD increases.

Here, we developed a stool host RNA-seq protocol that detects more protein-coding genes than previously reported methods. Using it, we generated 285 RNA-seq libraries from four clinical groups (normal, IBD, CRC, and APL) to address four questions. First, which cells do the mRNA transcripts in stool come from? Second, can adaptive immune signals, particularly T-cell and B-cell receptor (TCR and BCR) sequences, be detected in stool host RNA? Third, can stool mRNA distinguish between different GI diseases, specifically CRC and IBD? Fourth, can stool mRNA capture changes in immune cell populations across IBD severity, and so serve as a biomarker for disease monitoring? We find that stool host RNA is a census of shed cells, comprising epithelial cells from along the gastrointestinal tract as well as innate and adaptive immune cells, and that it also captures TCR and BCR repertoires. We further found that CRC and IBD leave distinct, separable transcriptional programs in this census, and that within IBD, neutrophil influx, the immune cell fraction, and the adaptive immune response all scale with disease severity and can be read from a single library.

## Results

### The QC metrics of stool host transcriptome sequencing

We developed an RNA-seq protocol that efficiently isolates stool mRNA and converts it into an Illumina sequencing library (Fig. 1a). In brief, oligo(dT) probes capture the human mRNA specifically and leave the abundant microbial nucleic acids behind. The protocol uses an optimized lysis buffer and conditions that release cellular RNA from human cells efficiently while protecting the mRNA from further degradation. The oligo(dT) probes capture both the relatively intact cellular RNA and the more degraded cell-free RNA, together with optimized mRNA binding and elution conditions and a workflow that can easily scale with stool input. Together, these improvements raised the quality of stool RNA-seq. Compared with existing methods for profiling the stool host transcriptome, ours detects far more protein-coding genes: the median across the 285 libraries was 6,300 (Fig. 1b), almost double the largest number previously reported, even under a stringent definition (genes with ≥ 10 reads, after removing reads with mapping quality (MAPQ) < 30). In RNA-seq libraries from low-biomass samples such as plasma, urine and stool, a high number of detected genes can also be an artifact of incompletely digested residual DNA. As a check, the median intergenic rate of the reads mapped to the human genome was only 0.7%, indicating very little DNA contamination. The mapping rates of these libraries were comparatively high, with a median of 49%, and library complexity was high for a low-input sample type, with a median of 1.7 million unique CDS-exon tags (Fig. 1b).

**Figure 1.**
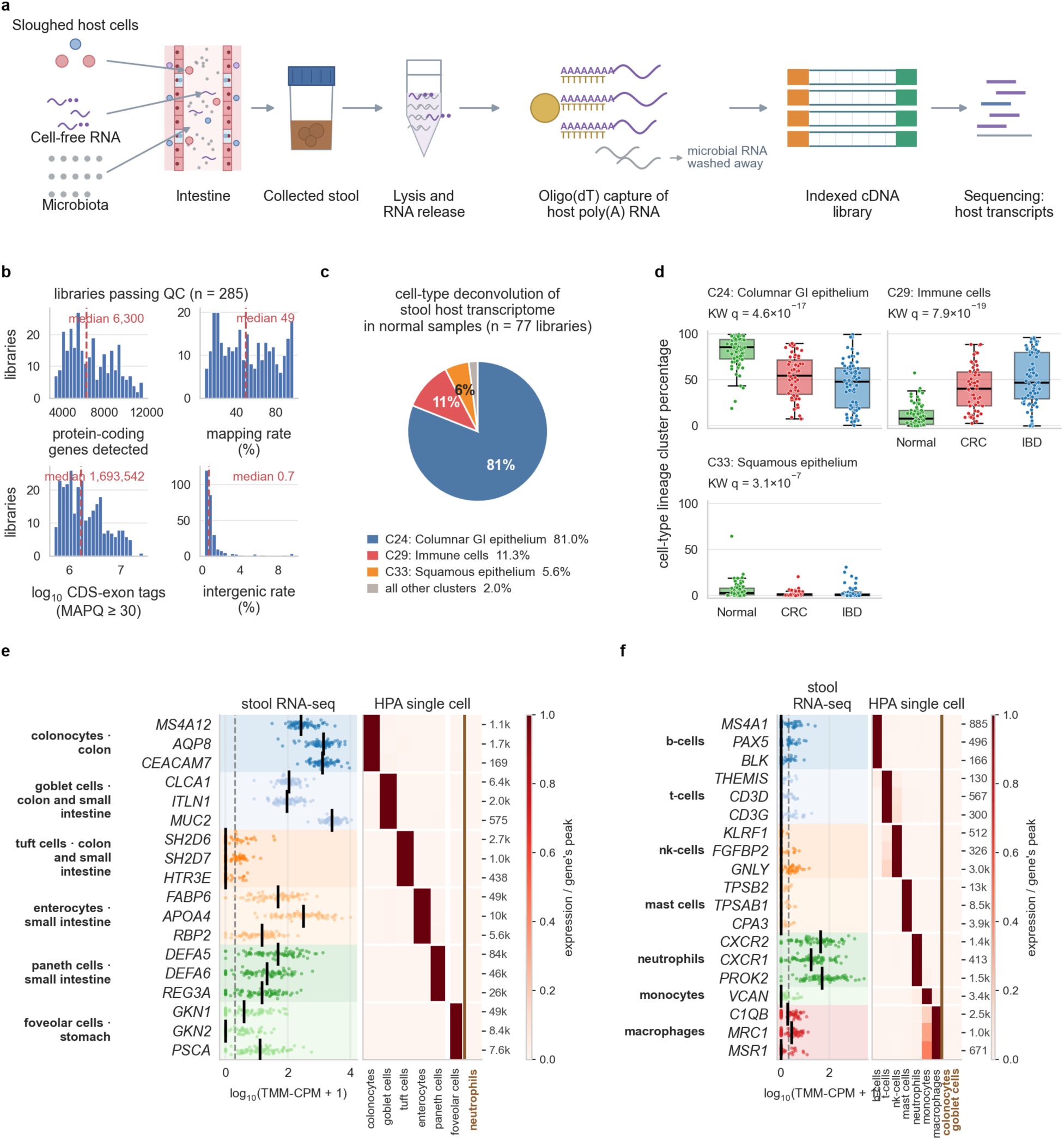
Stool host RNA is a census of the cells entering the lumen. **a**, The assay workflow of the stool host transcriptome sequencing. Host cells shed into the lumen, which is dominated by gut microbiota, and both are present in the collected stool. Cell lysis releases the cellular RNA; oligo(dT) beads capture the host poly(A) transcripts by their tails and wash the microbial RNA away; the retained RNA becomes one indexed cDNA library per stool, sequenced to reads that are aligned to the host genome. **b**, Quality metrics of the 285 libraries in this study. **c**, The cell-type decomposition of the stool host transcriptome using cell-type deconvolution. The pie chart shows the mean percentage of cell-type lineage clusters in normal samples (n = 77); clusters reaching 5% in any clinical group (Normal, CRC, or IBD) are named; the remaining 13 are pooled and named as all other clusters. **d**, The percentage of the cell type lineage clusters (from cell-type deconvolution) across stool host transcriptome in Normal (n = 77), CRC (n = 55) and IBD (n = 76); q, Kruskal–Wallis (KW) over the three groups, Benjamini–Hochberg corrected. **e**, **f**, Cell-type-specific marker panels in Normal stool for the columnar gastrointestinal epithelium along the tract, from colon to stomach (**e**), and for the major immune cell types (**f**). Left, the gene expression level (in log₁₀(TMM-CPM + 1)) of the cell-type specific markers in stool host transcriptome of normal samples. The dashed line marked the median expression level. Right, each marker gene’s expression across the single-cell RNA-seq reference data (HPA). Every value in a row is divided by that gene’s highest value (peak value) among all the cell types in the panel. The peak value for each gene is printed at the right of each row. Columns to the right of the vertical rule (brown) are control cell types that dominate stool libraries: neutrophils for the epithelial panel, and colonocytes and goblet cells for the immune panel.

### Stool host RNA is a census of the cells entering the lumen

With this unbiased profiling method, we prepared 285 stool host RNA-seq libraries from four clinical groups: Normal (n = 77), APL (n =63), CRC (n=55), and IBD (n = 90) (Methods, Supplementary Fig. 3). The CRC group has 7 stage I, 17 stage II, 16 stage III, 14 stage IV, and 1 unknown-stage sample. The IBD group has 42 ulcerative colitis (UC) and 46 Crohn’s disease (CD) samples, and 2 of unrecorded subtype. We first asked which cell types the stool host mRNA originates from and in what proportions, because this determines which diseases, and at which sites, a stool mRNA test could surveil. We used cell-type deconvolution to estimate the share of transcripts derived from each cell type. A common pitfall of deconvolution is that cell types of the same lineage have very similar expression profiles, so their individual shares cannot be assigned accurately. To avoid this, we first grouped the cell types of the single-cell reference into lineage clusters and deconvolved at the cluster level (Methods; Supplementary Table 1). In people without gastrointestinal disease (the Normal group), on average 81% of stool mRNA transcripts derived from columnar gastrointestinal epithelium, 11% from immune cells and 5.6% from squamous epithelium, most likely shed from the anal canal (Fig. 1c). In the CRC (n = 55) and IBD (n = 76, removed 14 remission samples) groups the share of immune-cell transcripts rose markedly, to a mean of 41% in CRC and 45% in IBD (Fig. 1d).

Deconvolution reports lineages, not cell types. To ask which cell types are detectable in stool, we examined the expression of signature genes for the major cell types of the columnar gastrointestinal-epithelium cluster and of the immune cluster (Fig. 1e, f); how these genes were selected is described in Methods and Supplementary Fig. 6. In the reference single-cell RNA-seq dataset, each of these genes is highly specific to its cell type among the major members of both clusters. A marker must also be nearly absent from the cell types that dominate stool (neutrophils, colonocytes and goblet cells), unless it marks one of them, or it could not indicate the presence of a rarer cell type; the reference panels of Fig. 1e, f show that each marker is indeed low in those abundant cell types. Transcripts from both the lower and the upper gastrointestinal tract were detectable, with colonocytes and goblet cells the dominant sources. Tuft-cell signature genes were barely detectable, as expected for a cell type that is rare in the healthy colon ^12^, and the abundance of epithelial transcripts tended to fall with the distance of the organ of origin from the anus. In the immune cluster, the most abundant signature genes were those of neutrophils, which are routinely delivered to the lumen to meet pathogens before they invade, whereas the signature genes of adaptive immune cells were at very low levels in the Normal group, which serves as a clean baseline for the disease samples.

### An adaptive-immune repertoire comes with the census

We next asked whether adaptive immune cells become more abundant in gastrointestinal disease. The B-cell/plasma-cell transcript *JCHAIN*, which encodes the small polypeptide that polymerizes immunoglobulins ^13^, was higher in CRC and IBD than in Normal stool, and so was the T-cell transcript *CD3E* which encodes a subunit of the T-cell receptor CD3 complex (Fig. 2b); the curated B-cell and T-cell signature scores show the same ordering (Supplementary Fig. 7). More interestingly, T-cell receptor (TCR) and B-cell receptor (BCR) sequences could be assembled from the stool RNA-seq reads themselves (Fig. 2a). Productive BCR clonotype counts correlated with *JCHAIN* expression (Spearman ρ = 0.68) and productive TCR clonotype counts with CD3E (ρ = 0.52) across the 208 (Normal: 77, CRC: 55, IBD: 76, the 14 IBD remission samples were removed) libraries of Fig. 2a, b (Supplementary Fig. 2). Of the 5,409 productive clonotypes recovered from the 285 libraries, 20 TCR and 606 BCR light-chain CDR3 sequences matched entries in public databases (VDJdb, IEDB and CoV-AbDab), with the hits concentrated on common human exposures—cytomegalovirus, SARS-CoV-2, influenza and Epstein–Barr virus (Fig. 2c). Such matches are evidence that an assembled sequence has been observed independently, not antigen-specificity calls: immunoglobulin light-chain CDR3s in particular are short and frequently shared, and the databases are heavily weighted toward SARS-CoV-2 because the database holds far more SARS-CoV-2 records than records for any other antigen. In addition, 7 of 88 T Cell Receptor Alpha chain (TRA) clonotypes carried mucosal-associated invariant T (MAIT)-like α chains (*TRAJ33* or *TRAJ20* junctions, five of them on the canonical *TRAV1-2*; Fig. 2d). MAIT cells recognize riboflavin (vitamin B2) metabolites synthesized by gut microbes ^14^, so their presence supports the conclusion that stool host RNA-seq recovers TCR sequences that genuinely belong to the gut immune system.

**Figure 2.**
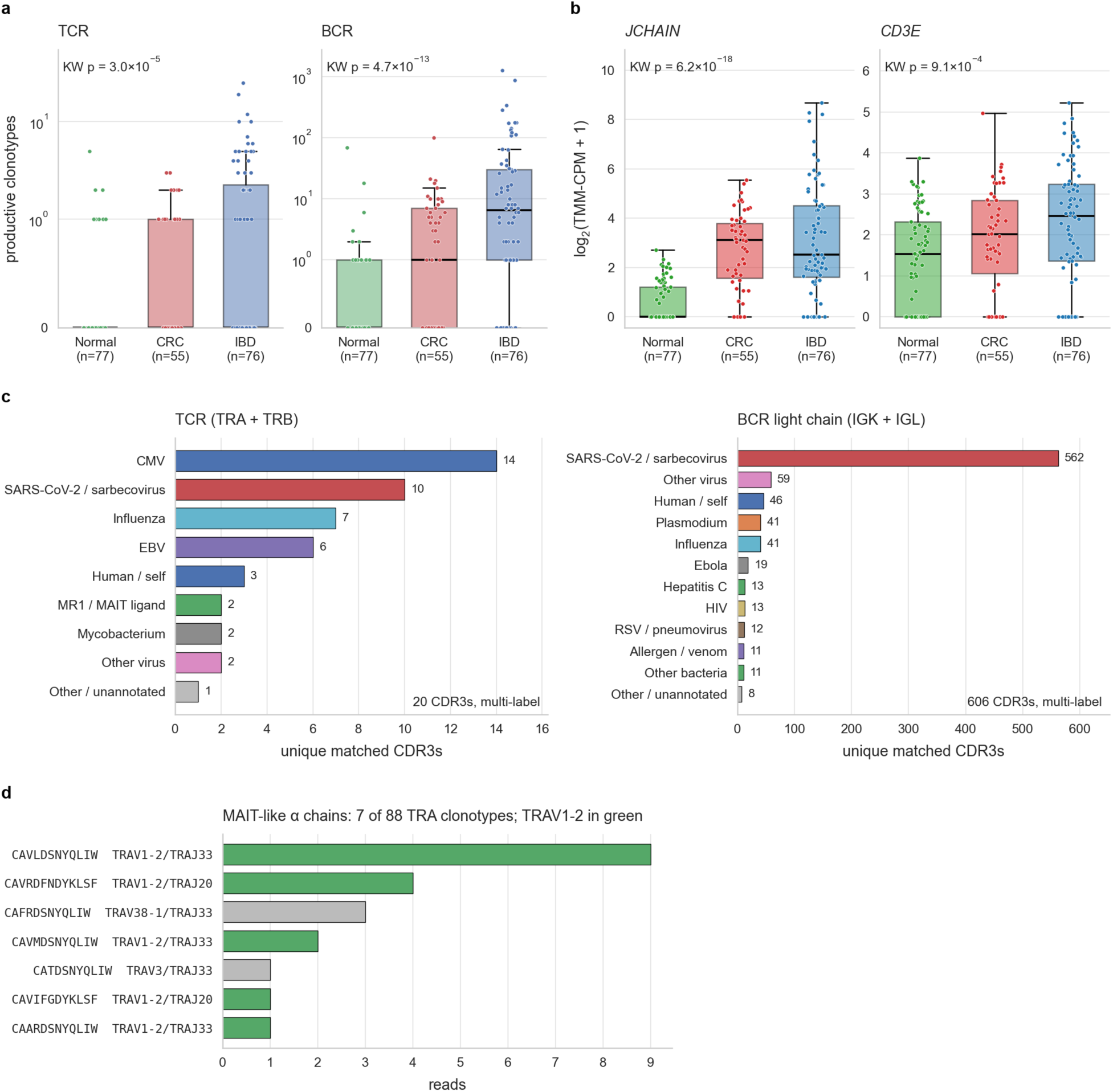
An adaptive-immune repertoire comes with the census. **a**, Productive TCR and BCR clonotypes, assembled by TRUST4 using the stool RNA-seq data, in Normal (n = 77), CRC (n = 55) and IBD (n = 76; endoscopic remission set aside, the same libraries as Fig. 3a) libraries; KW, Kruskal–Wallis test shows the statistical significance over the three groups. **b**, Expression of the B-cell & plasma cell specific gene *JCHAIN* and the T-cell specific gene *CD3E* in the same libraries as in **a**, as log₂(TMM-CPM + 1); KW, as in **a**. **c**, What the assembled TCR and BCR receptor sequences from all the stool RNA-seq libraries match in public databases (VDJdb, IEDB and CoV-AbDab). The CDR3 sequences were counted as a match only when its CDR3 core was identical to a record from the same locus. Bars give the number of matched CDR3s in each antigen class, for TCRs (TRA + TRB, left) and BCR light chains (IGK + IGL, right); the total number of matched CDR3s is printed in each plot. A CDR3 annotated to several antigens is counted once in each class, so the bars add up to more than that total. **d**, MAIT-like α chains among the 88 TRA clonotypes of the same libraries: CDR3α, V/J usage and supporting reads; green, canonical *TRAV1-2*.

### Stool host mRNA distinguishes gastrointestinal diseases

Can stool mRNA distinguish gastrointestinal diseases from one another, or does it only report how inflamed the gut is? To answer this we performed principal-component analysis on the 2,000 most variable genes, setting aside the IBD libraries in endoscopic remission (n = 14) because a healed mucosa is expected to resemble Normal stool. Plotting component scores by disease group, the dominant axis (PC1, 62% of variance) separated both CRC and IBD from Normal stool (single-component AUC 0.90 and 0.91). The component that best separated CRC from IBD was PC6 (AUC 0.86); no other component reached 0.7. Plotting PC1 against PC6 shows the separation of Normal, active IBD, and CRC libraries (Fig. 3a): this unsupervised decomposition already separates the libraries by diagnosis, which indicates that the signals distinguishing active IBD from CRC are strong. We then ran pathway enrichment on the 100 genes loading most positively and most negatively on each of PC1 and PC6 (Enrichr; GO Biological Process, KEGG, Reactome and MSigDB Hallmark, with the 2,000 genes of the decomposition as background; Fig. 3d). The positive end of PC1 was enriched for neutrophil degranulation and innate immunity, as expected given that neutrophil-derived mRNA dominates the immune signal in the stool host transcriptome. Coloring the PCA by a neutrophil score (the mean z-score of canonical neutrophil-specific genes, Methods) showed that this score separates Normal from IBD and CRC libraries along PC1 (Fig. 3b), with which it correlated almost perfectly (Spearman ρ = 0.99). The positive end of PC6, toward which IBD lies, was enriched for the interferon-γ response, and its negative end, toward CRC, for epithelial– mesenchymal transition (EMT) and TNF-α/NF-κB signaling (Fig. 3d). EMT is a hallmark of cancer pathogenesis, and interferon and TNF-α signaling, although strongly up-regulated in IBD, are also involved in CRC. To characterize the host mRNA signals that separate IBD from CRC more fully, we performed differential expression analysis (using DESeq2) between the 76 active IBD and 55 CRC libraries and identified 522 genes higher in CRC and 769 higher in IBD (Fig. 3e). On the volcano plot we labeled the differentially expressed genes belonging to four canonical programs directly linked to IBD and CRC pathogenesis (WNT signaling, EMT/stroma, proliferation/E2F and the IFN-γ response). We also calculated the mean z-score of the genes in these four programs by clinical diagnosis groups (Fig. 3f). Consistent with the biology of the two diseases, the IFN-γ response program was higher in IBD than in CRC and the other three were higher in CRC than in IBD. Notably, CRC libraries also scored higher than Normal on the IFN-γ program, consistent with the inflammatory component of CRC pathogenesis.

**Figure 3.**
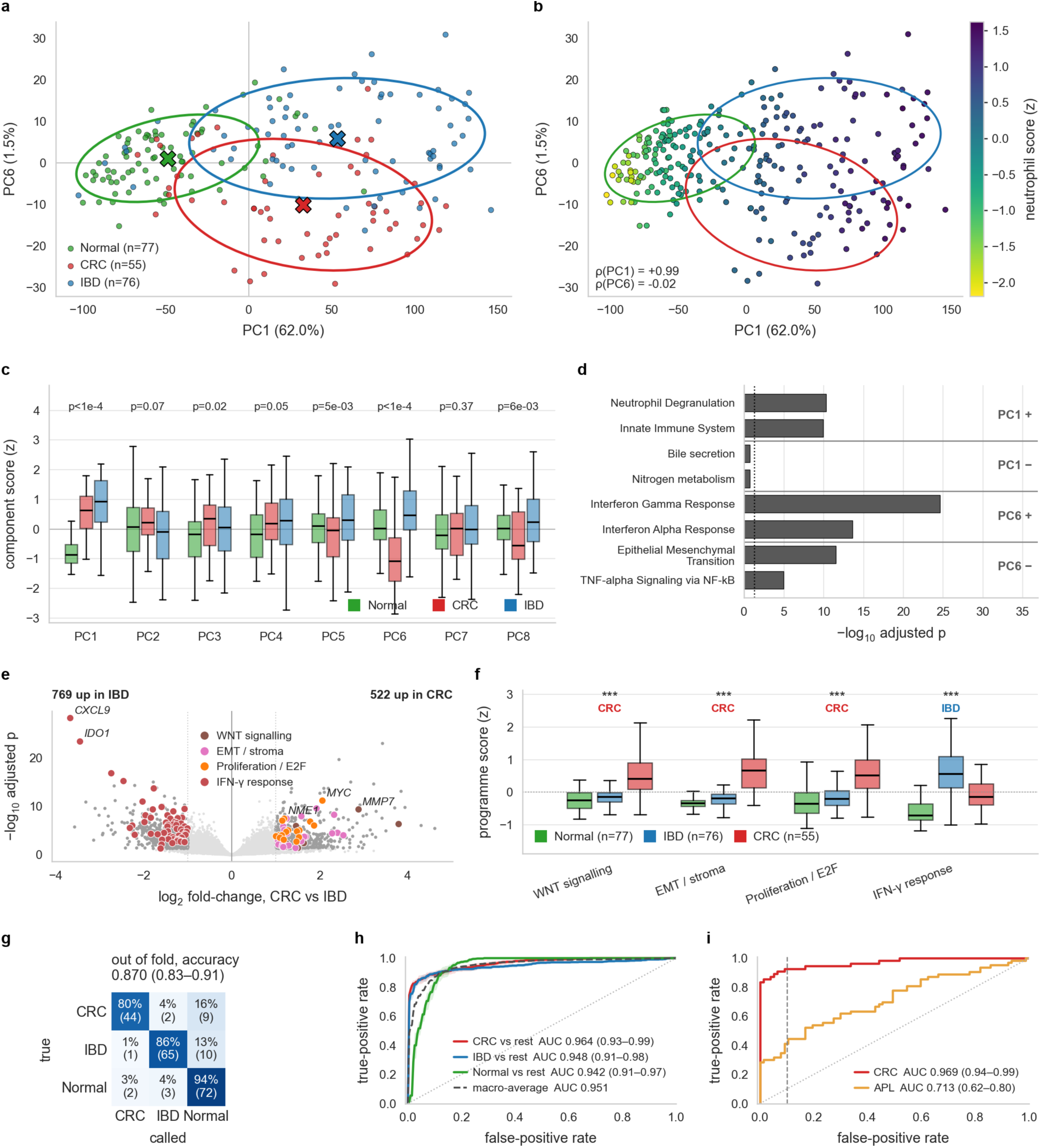
Colorectal cancer and IBD write distinct programs into the census. Throughout: Normal n = 77, CRC n = 55, IBD n = 76, remission excluded. **a**, PCA of the 271 libraries on their 2,000 most variable genes, drawn on PC1 and PC6, the component that best separates CRC from IBD (single-component AUC 0.86). APL (n = 63) is in the decomposition but not drawn. Crosses, group means; ellipses, 68% contours (1.51 s.d.). **b**, The same plane, each library colored by its 15-gene neutrophil score, with the contours of **a**; ρ, Spearman correlation of the score with each component (+0.99 with PC1, −0.02 with PC6). **c**, Every component of the decomposition, z-scored, by group; p, Kruskal–Wallis, unadjusted. **d**, Enrichment of the 100 genes loading most positively and the 100 most negatively on PC1 and PC6, against the 2,000 genes of the decomposition as background (Methods); the two most significant terms per pole are shown. **e**, Differential expression between CRC and IBD (DESeq2). At adjusted p < 0.05 and |log₂ fold-change| > 1, 522 genes are higher in CRC and 769 in IBD. Colored genes belong to four canonical programs: Wnt signaling, epithelial–mesenchymal transition/stroma and proliferation/E2F on the CRC side, interferon-γ response on the IBD side. **f**, Scores of those programs (mean z-score of the member genes) by group. Stars, CRC versus IBD, two-sided Mann–Whitney U, unadjusted (*, p < 0.05; **, p < 0.01; ***, p < 0.001); the label names the higher group. **g**, **h**, A three-class logistic-regression model on the whole transcriptome (Methods). **g**, Confusion matrix of each library’s consensus out-of-fold call, row-normalized with counts in brackets; title, accuracy of those calls with its 95% bootstrap CI over libraries. **h**, One-versus-rest ROC curves, mean over the ten assignments with ± 1 s.d. bands; AUC as in **g**. **i**, A two-class model of CRC and APL pooled (n = 118) against Normal, read as CRC versus Normal (AUC 0.969, 0.94–0.99) and as APL versus Normal (AUC 0.713, 0.62–0.80). Dashed line, 90% specificity, at which 0.90 of cancers and 0.42 of advanced adenomas are called.

Distinguishing IBD, CRC and Normal stool has direct clinical utility: for patients with mild gastrointestinal symptoms, a non-invasive stool test that flags likely IBD or CRC could triage them to colonoscopy and avoid weeks or months of waiting. We therefore built a three-class multinomial logistic-regression model on 208 libraries (CRC 55, IBD 76, Normal 77; Methods). Under five-fold cross-validation repeated ten times, each library was given its consensus out-of-fold call, the class it was assigned most often; the accuracy of these calls was 0.870 (95% bootstrap confidence interval 0.83–0.91), with one-versus-rest AUCs of 0.964 (0.93–0.99), 0.948 (0.91–0.98) and 0.942 (0.91–0.97) for CRC, IBD and Normal respectively (macro-average 0.951; Fig. 3g, h). The errors were mostly disease libraries called Normal—16% of CRC and 13% of IBD—and these were predominantly early-stage cancers and mild colitis: of the nine CRC libraries called Normal, seven were stage I or II and none was stage IV, and of the ten IBD libraries, seven were mild.

### Advanced neoplasia can be called from the stool transcriptome alone

Stool-based screening for CRC and APL is among the most widely used non-invasive stool tests. We therefore asked whether a classifier built directly on the stool host transcriptome can detect CRC and APL. We trained a two-class logistic-regression model on the 20 genes with the largest Welch *t*-statistic within each training fold, using five-fold cross-validation and CRC (n = 55), APL (n = 63), and Normal (n = 77) samples (Methods). It separated CRC from Normal with AUC 0.969 (95% bootstrap confidence interval 0.94–0.99) and APL from Normal with AUC 0.713 (0.62–0.80). The sensitivity is 90% for CRC and 42% for advanced adenomas at 90% specificity (Fig. 3i). The genes chosen in at least 20 of the 25 training folds (five folds × five repeats) were *HBB*, *HBA1*, *HBA2*, *TGFBI*, *SPARC*, *MMP1*, *MMP3*, *MMP7*, *IL6*, *IL33* and *TUBB3*. *HBB*, *HBA1* and *HBA2* encode hemoglobin, whose protein is the analyte of the fecal immunochemical test (FIT) and of the combined DNA-methylation and host-mRNA stool tests used for CRC screening ^15,16^. *MMP3*, *MMP7*, *TUBB3* and *TGFBI* have been previously reported as stool-based CRC biomarkers from tissue RNA-seq and validated by stool RT-PCR ^17^. Even with this modest sample size, therefore, stool RNA-seq identified credible biomarkers for CRC screening. Compared with the conventional strategy of predicting fecal biomarkers from tissue data and validating them in stool by PCR or small targeted panels, stool whole-transcriptome sequencing offers a faster, higher-throughput way to discover fecal biomarkers for gastrointestinal diseases.

### Immune signals in the stool host transcriptome track the severity of IBD

Better ways to monitor disease activity in IBD are an unmet clinical need. The standard is endoscopy, and the only non-invasive alternative in routine use is the fecal calprotectin immunoassay, whose accuracy is limited, especially in Crohn’s disease ^4^. We therefore asked whether stool host RNA can monitor IBD activity with better accuracy. We took the 66 IBD libraries with an endoscopic grade evaluated within 30 days of stool collection and a recorded subtype—33 ulcerative colitis (UC) and 33 Crohn’s disease (CD); remission 12, mild 17, moderate 17, severe 20—together with 24 healthy samples as a reference band. The healthy samples are a subset of the Normal group that, in addition to a negative colonoscopy, showed no evidence of inflammation or other lesions (Methods). We first examined the two genes encoding the calprotectin complex, *S100A8* and *S100A9*, since fecal calprotectin is well established to rise during IBD flares. Across the 66 libraries, both correlated positively with endoscopic grade (Spearman correlation coefficient ρ = 0.66 and 0.65; p ≤ 2.8 × 10⁻⁹; Fig. 4a). Calprotectin is used as an indicator of neutrophil abundance in the gut because of its high expression in neutrophils ^18^, but one protein, or two genes, may not represent the neutrophil load accurately. We therefore defined a neutrophil panel of 15 genes that are highly and specifically expressed in neutrophils and took the mean z-score of their expression as a neutrophil score (Methods). The score correlated with grade more closely than *S100A8* or *S100A9* alone (Spearman correlation coefficient ρ = 0.75) and separated each grade from the next with statistical significance (Fig. 4a; two-sided Mann–Whitney U, p = 0.012 for remission versus mild, 0.013 for mild versus moderate and 0.0011 for moderate versus severe). Beyond neutrophils, the immune-cell share of transcripts estimated by deconvolution (Methods) also rose with severity, reflecting the growing inflammatory infiltrate (Fig. 4a). Host RNA also tracks the lymphocytes: productive BCR and TCR clonotype counts rose with severity (Fig. 4b), as did the B- and T-cell signature scores (Fig. 4c).

**Figure 4.**
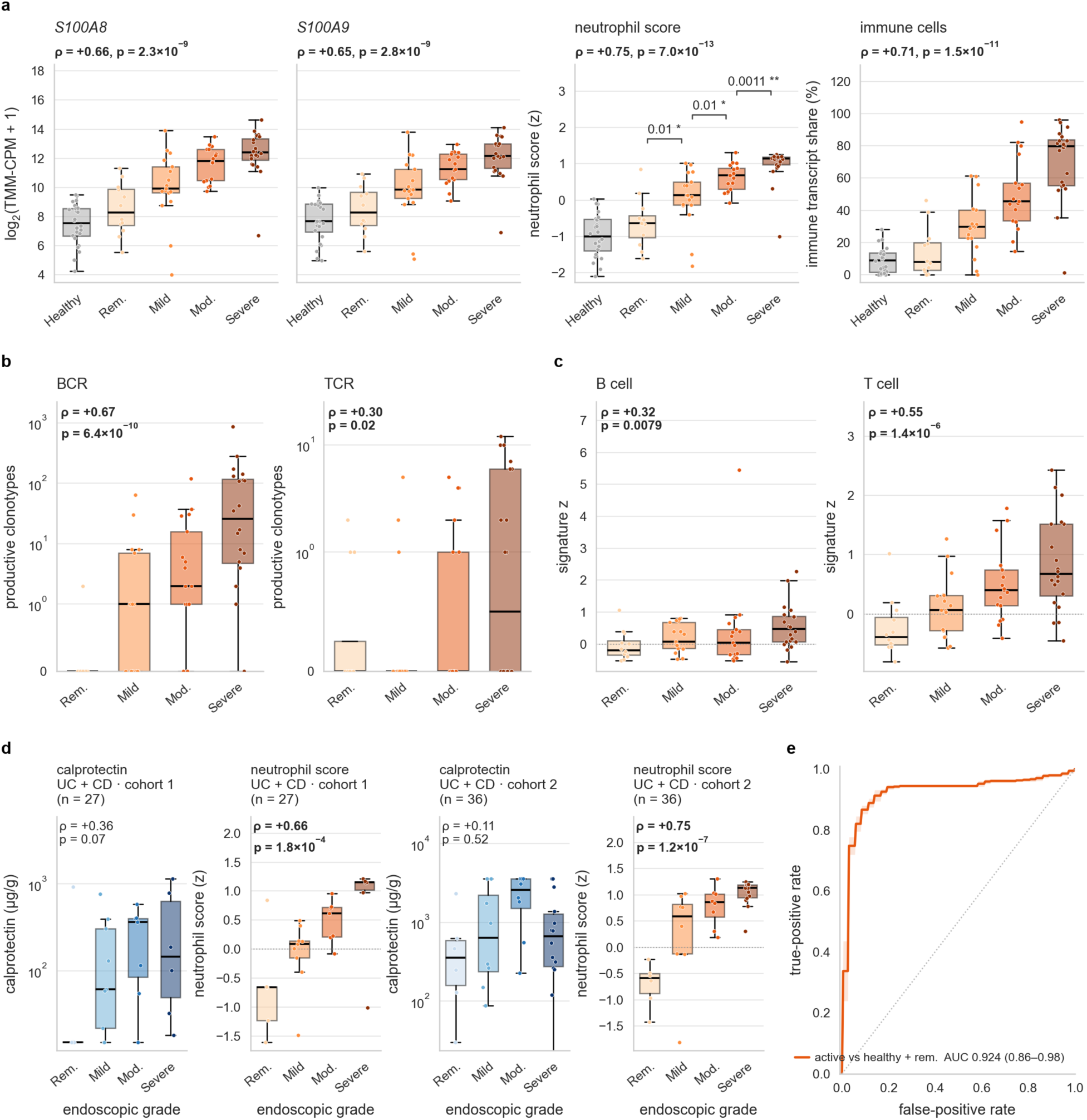
Stool host transcriptome tracks both the neutrophil influx and the adaptive-immune response across the severity grades of inflammatory bowel disease. **a**, *S100A8*, *S100A9*, the 15-gene neutrophil score and the immune-cell share (by cell-type deconvolution) of the library, each against endoscopic grade, in 24 healthy libraries and 66 IBD libraries (ulcerative colitis 33, Crohn’s disease 33; remission 12, mild 17, moderate 17, severe 20). ρ and p, Spearman correlation with grade over the IBD libraries. Brackets over the neutrophil score, two-sided Mann–Whitney U between neighboring grades (remission versus mild, mild versus moderate, moderate versus severe), unadjusted p; *, p < 0.05; **, p < 0.01. The two genes and the score are TMM-normalized, and the two genes share one axis; the immune-cell share is the immune-cell column of the deconvolution in Fig. 1c, d, as a percentage of transcripts. **b**, **c**, The adaptive-immune readouts against the same grades, in the same 66 IBD libraries as **a**: productive BCR and TCR clonotypes (**b**) and B-cell and T-cell signature scores (**c**); ρ and p, Spearman correlation with grade, unadjusted. **d**, Clinical fecal calprotectin compared with the 15-gene stool neutrophil score, each against the endoscopic grade band, in the IBD libraries of both subtypes that have a paired calprotectin measurement, split by recruiting cohort with different calprotectin reagents; ρ and p, Spearman. **e**, A classifier of active disease from the 15-gene neutrophil score: active disease (mild, moderate or severe, n = 54) against the quiescent gut, the 24 healthy and 12 remission libraries together. AUC, mean over assignments with its 95% bootstrap confidence interval over libraries resampled within grade, 0.924 (0.86–0.98).

To demonstrate the potential clinical utility, we compared the stool-mRNA panel with the conventional method, fecal calprotectin. Because the number of IBD libraries in each endoscopic grade is small, a machine-learning model trained on this dataset would risk wide confidence intervals and over-fitting, so we took a biology-driven approach and used the neutrophil score directly as the stool-mRNA alternative to calprotectin. The IBD libraries were split into two cohorts by the recruiting hospital, whose calprotectin reagents and instruments differ (Methods). The stool-RNA neutrophil score tracked endoscopic grade more closely than calprotectin did on the identical samples, in both recruiting cohorts and in both subtypes (Fig. 4d; Supplementary Fig. 5): in UC, the Spearman correlation with endoscopic grade was ρ = 0.71 for the neutrophil score against 0.48 for calprotectin in cohort 1 (n = 12) and 0.68 against 0.39 in cohort 2 (n = 20); in CD, 0.53 against 0.19 in cohort 1 (n = 15) and 0.76 against 0.62 in cohort 2 (n = 16); and pooled across subtypes, 0.66 against 0.36 in cohort 1 (n = 27) and 0.75 against 0.11 in cohort 2 (n = 36; Fig. 4d). CD in cohort 1 is the weakest cell: calprotectin did not track grade there (p = 0.49), although the stool score did (p = 0.044). Finally, a classifier on the neutrophil score as its single feature separated active IBD from the quiescent gut—IBD in remission and healthy volunteers together—with an out-of-fold AUC of 0.924 (Fig. 4e).

## Discussion

Everything reported here is enabled by a stool host RNA-seq method that surveys the stool host transcriptome without targeting particular genes and that detects more human genes than any previously reported approach. The first finding is a systematic account of the cells of origin of host RNA in stool, something never done systematically or unbiasedly before. Normal stool is dominated by intestinal epithelium, whose colonocyte, goblet, enterocyte, Paneth and gastric markers are each individually recoverable and specific against a single-cell reference; the immune fraction is small and neutrophil-led; and lymphocyte transcripts sit near the detection floor.

Disease inverts the mixture toward immune cells, and the adaptive compartment becomes readable not only as transcripts but as assembled antigen receptors: these track their lineage genes, match receptors recorded in public databases and include the MAIT-like α chains expected at a mucosal surface. To our knowledge, this is the first recovery of TCR and BCR sequences from stool host RNA.

We demonstrated three potential clinical applications of stool host transcriptome sequencing. The first is differential diagnosis. CRC, IBD and Normal stool carry distinct transcriptional signatures, and the genes that separate cancer from colitis resolve into canonical disease programs: Wnt signaling, stroma and proliferation on the cancer side, and an interferon-γ response on the colitis side. A three-class model read on these features separated CRC, IBD and Normal stool with one-versus-rest AUCs of 0.94 to 0.96, which shows that stool host RNA carries the identity of the disease and not only the presence of inflammation. To our knowledge, this is the first study showing that the stool host transcriptome can discriminate between different gastrointestinal diseases.

The second application is screening for advanced neoplasia. A two-class model distinguished CRC from Normal at an AUC of 0.97; APL from Normal at 0.71, and it did so on hemoglobin, stromal and inflammatory transcripts rather than on the lesion’s own epithelial program. The census reports what a lesion sheds, and an adenoma sheds much less than CRC.

The third application is monitoring IBD activity. A classifier on the 15-gene neutrophil score alone separated active disease from the quiescent gut at an AUC of 0.92, and the same libraries reported the infiltrate behind that score: the immune-cell share of transcripts, the neutrophil score and the B-cell and T-cell signature scores all rose with endoscopic grade, as did the number of productive BCR and TCR clonotypes. The stool neutrophil score also tracked endoscopic grade more closely than the conventional fecal calprotectin measured on the same stool, in both recruiting cohorts and in both subtypes. The correlation was weakest in Crohn’s disease in cohort 1, as expected for a patchy transmural disease sampled from the lumen, and should be read as a boundary of the approach rather than a failure of the assay.

Several limitations apply. The activity cohort is modest: 66 graded IBD libraries, 63 of them with a paired calprotectin. Its grade axis pools UC and CD, which were graded with different instruments—the Mayo endoscopic subscore and SES-CD—that score different lesions and were mapped onto a single four-level band. The subtypes also differ in extent, since UC is confined to the colon whereas CD usually involves the ileum and can involve any segment of the gut, so the endoscopic reference may not capture all the disease a stool sample integrates. We pooled them because the number of libraries within each subtype is small, and the two are unevenly distributed across grades (19 of 33 UC libraries severe, 1 of 33 CD). Detection of a marker is evidence that transcripts of that cell type are present, not a cell count, and the deconvolution reports shares of transcripts, which depend on the mRNA content of each lineage.

In this proof-of-principle study we have shown that stool host transcriptome sequencing recovers mRNA not only from the gut epithelium but also from immune cells, together with their antigen-receptor repertoire, and we have demonstrated its clinical potential in gastrointestinal differential diagnosis, CRC screening and the monitoring of IBD activity. The same readout could serve diagnosis, prediction of drug response and disease monitoring across a wider range of gastrointestinal conditions, such as *Clostridioides difficile* infection and irritable bowel syndrome.

## Methods

### Ethical review

The Medical Ethics Committee of Renji Hospital, School of Medicine, Shanghai Jiao Tong University (KY2025-108-B), Shenzhen Hospital of Southern Medical University (approval no. NYSZYYEC2026K136R001), Huizhou First Hospital (KYLL-2025-044-04) and Jiangmen Central Hospital (approval no. [2024] 228 号 D) gave ethical approval for this work.

### Study participants

The stool samples in this study were collected from Renji Hospital, School of Medicine, Shanghai Jiao Tong University, Shenzhen Hospital of Southern Medical University, and a contract research organization (ZunTai Medical Technology, Guangzhou, China).

On the basis of colonoscopy, Normal participants were further categorized as healthy (colonoscopy-negative with no inflammation or lesion), NEG (colonoscopy-negative with hyperplastic polyps or a non-neoplastic lesion) or non-advanced precancerous lesion (NAPL: tubular adenoma(s) < 10 mm in diameter, serrated adenomatous polyp(s) < 10 mm, or ≥ 20 hyperplastic polyps).

IBD subtype (UC or CD), Montreal classification, disease location, endoscopic score and date, and fecal calprotectin concentration were obtained from clinical records. Endoscopic activity was recorded as the Mayo endoscopic subscore for UC and as an SES-CD category for CD, and both were mapped onto a common four-level grade (remission < mild < moderate < severe). IBD libraries in endoscopic remission (n = 14) were excluded from every between-diagnosis comparison—cluster shares (Fig. 1d), repertoire size and lineage transcripts (Fig. 2a, b), principal-component analysis, differential expression, program scores and the diagnostic classifiers (Fig. 3)—because a healed mucosa is expected to shed little disease-specific material. They were retained in the antigen-database and invariant-chain analyses (Fig. 2c, d) and in the IBD disease activity analyses (Fig. 4), in which remission is one of the grades and, with the healthy libraries, forms the negative class of the activity classifier (Fig. 4e). The activity-monitoring set comprised IBD libraries with an endoscopic grade and an endoscopy dated within 30 days of stool collection, in either direction. Of the 90 IBD libraries, 16 had no recorded grade and 6 were graded but outside the 30-day window (no graded library lacked an endoscopy date), leaving 68; the 2 of these without a recorded IBD subtype were also excluded, leaving 66 (UC 33, CD 33; Supplementary Fig. 3).

### Stool sample collection, host RNA enrichment, library preparation and sequencing

Fresh stool samples were collected by patients in fecal collection tubes, delivered to the hospital within 4 h, and stored at −80 °C. Human RNA was isolated from 1 g of stool per sample. The method first lyses human cells in lysis buffer and enriches the human mRNA on oligo(dT) beads.

We quantified the extracted RNA by NanoDrop (concentration, A260/A280, and A260/A230), measured host RNA content by GAPDH amount using RT-qPCR (Takara OneStep Prime Script III RT-qPCR 2X Master mix (Takara Bio, part #RR601)), and stored it at −80 °C. The library-preparation procedure specifies a total RNA concentration ≥100 ng µl⁻¹ (NanoDrop One C, Thermo Fisher Scientific) and a *GAPDH* RT-qPCR Ct ≤31 as its input requirement for the downstream library preparation.

The isolated fecal RNA was converted into strand-specific, dual-indexed libraries with the Fast RNA-seq Lib Prep Kit V2 (ABclonal, China), using 16 PCR cycles and unique dual-index primers. Libraries were sequenced as 2 × 150 bp paired-end reads on a NovaSeq 6000 to a median of 47 million clean reads (range 25–173 million) after adapter and quality trimming.

### Fecal calprotectin measurement

In Cohort 1, the calprotectin concentration was measured using the Fecal Calprotectin Detection Kit (Guangzhou Forreal Biotechnology Co., Ltd.). In Cohort 2, the calprotectin concentration was measured using the BÜHLMANN fCAL ELISA (BUHLMANN, Switzerland).

### Read processing and per-library quality metrics

Reads were processed with the RNA-seq pipeline: trimmed and quality-filtered with fastp 1.3.0^19^ (default settings) and aligned to GRCh38 with STAR 2.7.11b^20^. Specificity was then imposed by restricting to records with MAPQ ≥ 30, both mates on the same reference and an absolute insert size ≤ 2,000 bp. Duplicates were marked on the filtered BAM files with Picard 3.4.0^21^ MarkDuplicates, and insert-size metrics were collected with Picard CollectInsertSizeMetrics on non-duplicate pairs.

Gene-level fragment counts were obtained with featureCounts^22^ (Subread 2.1.1), and the deduplicated matrix was used for all analyses. Mitochondrially encoded genes (MT-) were removed by the pipeline before its count matrices were written, so the deduplicated matrix contains no mitochondrial genes and every per-million denominator below refers to nuclear genes. The per-library QC metrics were: clean library size (reads after fastp); mapping rate and nuclear mapping rate (the fraction of clean reads aligned to the genome and, at MAPQ ≥ 30, to the nuclear genome); duplication rate (the fraction of MAPQ ≥ 30 mapped reads flagged as duplicates); CDS-exon tags (fragments overlapping coding exons at MAPQ ≥ 30); exonic and intergenic rates (the fractions of assigned MAPQ ≥ 30 tags in UTR and coding exons, and in intergenic regions); and detected protein-coding genes (genes with ≥ 10 deduplicated counts at MAPQ ≥ 30; Fig. 1b). Libraries with an intergenic rate > 10% or < 0.5 × 10⁶ CDS-exon tags were excluded. Both criteria are alignment metrics fixed before any expression analysis, so the filter is independent of the expression patterns analyzed downstream. The 285 libraries (healthy 24, NEG 33, NAPL 20, APL 63, CRC 55 and IBD 90) in this study already passed these filters.

### Normalization

One expression matrix serves every analysis. Counts per million (CPM) were recomputed from the deduplicated counts and corrected for composition bias with the trimmed mean of M-values (TMM)^23^ method, implemented in Python as a port of edgeR’s^24^ calcNormFactors with its default settings and fitted once on the 285 QC-passed libraries. This TMM-normalized CPM (TMM-CPM) is the unit of the deconvolution and the marker panels (Fig. 1c–f), the lineage transcripts and signature scores (Fig. 2b, Fig. 3b, Fig. 4a, c; Supplementary Figs. 2 and 7), the principal-component analysis, its pole enrichments and the diagnostic classifiers (Fig. 3a–d, g–i; Supplementary Fig. 9), the program scores (Fig. 3f; Supplementary Fig. 8) and the activity analyses (Fig. 4a, d, e; Supplementary Figs. 4 and 5). Differential expression (Fig. 3e) used raw counts. Normalized CPM (nCPM) appears only as the unit of the Human Protein Atlas (HPA) single-cell reference^25^, against which the marker and deconvolution criteria below are stated. All log transforms are log₂(x + 1) unless otherwise stated.

### Reference atlas and lineage clusters

The HPA single-cell type atlas^25^ (version 25.1, Ensembl 109; 154 cell types; nCPM) was restricted to genes reaching ≥1 nCPM in at least one cell type, and the 5,000 genes with the highest variance of log(nCPM + 1) were retained. Cell types were clustered by average-linkage hierarchical clustering on cosine distance and the tree was cut at a distance of 0.15 (33 clusters). A predefined list of twelve lineage merges, based on prior biological knowledge, was then applied in order, each joining the two clusters that held the named cell types and accepted only if the mean cosine distance between those clusters was ≤0.25 (all twelve were, at 0.16– 0.24): tuft cells with colonocytes; microglia with macrophages; platelets with megakaryocytes; erythrocytes with erythrocyte progenitors; late spermatids with undifferentiated spermatogonia, then oocytes with late spermatids, giving one germ-cell cluster; Müller glia with astrocytes, then retinal ganglion cells, astrocytes and retinal horizontal cells each with brain excitatory neurons, giving one neural–glial cluster; and myonuclei, then thymic myoid cells, with cardiomyocytes, giving one striated-muscle cluster. Six squamous cell types (esophageal apical, basal and suprabasal cells, basal and suprabasal keratinocytes, and ocular epithelial cells) were separated into a cluster of their own, because squamous cells reach stool by a different route from intestinal epithelium and pooling them with other epithelia obscures the dominant stool compartment. Hepatocytes were left as the singleton produced by the cut: because a cluster signature is the mean of its members, merging hepatocytes into the columnar gastrointestinal cluster would allow hepatocyte-restricted genes to score as markers of the compartment that dominates stool, and would leave no separate column to absorb apolipoprotein transcripts that both intestine and liver produce (see *Cluster-level deconvolution*). This procedure yielded 22 clusters (Supplementary Fig. 1; members in Supplementary Table 1).

### Cluster-level deconvolution

Cluster signatures were the mean nCPM of member cell types. Marker genes were selected at the cluster level on three criteria: (i) expression ≥10 nCPM in the highest-expressing cluster; (ii) that cluster accounting for ≥85% of the gene’s summed cluster means; and (iii) expression ≥10 nCPM in at least 34% of that cluster’s member cell types. Criterion (iii) ensures that a marker describes the cluster rather than one of its members: criteria (i) and (ii) score only the contrast between clusters, and because a cluster profile is a member mean, a gene expressed by a single member of a large cluster otherwise appears perfectly specific.

The resulting marker pool comprised 488 gene–cluster assignments, 479 of whose genes were present in the stool matrix. Clusters with fewer than five markers — the five singletons above and C22 (stromal and vascular cell types) — were excluded from the fit, leaving 16 of 22 clusters. Supplementary Table 1 lists every cluster with its signature genes, including the markers of the six clusters that were not fitted.

Each RNA-seq library (TMM-CPM) was deconvolved against the signature matrix, with each cluster’s profile scaled to unit Euclidean length so that clusters with very different absolute expression contribute on an equal footing, by dampened weighted least squares (DWLS)^26^: a non-negative least-squares (NNLS) initial estimate followed by four reweighted NNLS iterations with weights 1/ŷ², where ŷ is the fitted expression floored at the library’s 25th percentile of non-zero expression so that near-zero genes cannot dominate. This weighting minimizes relative rather than absolute error, which is appropriate when the mismatch between a tissue reference and a stool library is multiplicative. A fixed floor was used in place of the cross-validated dampening constant of the original method, which is selected for stability across gene subsets rather than for accuracy and did not improve any performance measure we examined.

The reported quantity is the transcript share: the fitted coefficients, renormalized to sum to one across clusters. Transcript share is not a cell-fraction estimate, because conversion to cell fractions requires equal mRNA content per cell, an assumption that does not hold across epithelial cells, neutrophils and erythrocytes. Fig. 1c shows the mean share in Normal libraries of each cluster reaching ≥5% mean share in Normal, CRC or IBD stool, with the remaining clusters pooled. Differences in the share of each of these clusters among Normal, CRC and IBD libraries (Fig. 1d) were tested by Kruskal–Wallis test with Benjamini–Hochberg correction across the clusters shown.

### Cell-of-origin marker panels

The panels of Fig. 1e, f ask which individual cell types can be distinguished within the two clusters that dominate stool. Thirteen cell types were specified in advance — colonocytes, goblet cells, tuft cells, enterocytes, Paneth cells and gastric foveolar cells from C24, and B cells, T cells, NK cells, mast cells, neutrophils, monocytes and macrophages from the immune cluster C29 — each with a curated panel of published markers from flow-cytometry gating, single-cell annotation and immunohistochemistry^27–34^, compiled from the literature rather than from these data. Intestinal panels follow the colonic^27^ and whole-gut^28^ single-cell atlases, the enterocyte and Paneth panels also the upper-gastrointestinal atlas^29^, the gastric foveolar panel that atlas^29^, and the hepatocyte panel the liver atlas^33^. Among the immune panels, B cells and monocytes follow the cross-tissue immune atlas^30^ and sorted-blood signatures^31^; T cells and NK cells the sorted-blood signatures^31^ and the T and NK clusters resolved within the colonic^27^ and liver^33^ atlases; mast cells the mast-cell cluster of the colonic atlas^27^; macrophages the macrophage clusters of the liver^33^ and whole-gut^28^ atlases; and neutrophils the sorted-blood signatures^31^ and CellMarker 2.0^32^. Classical flow-cytometry and immunohistochemistry markers compiled in PanglaoDB^34^ supplement every panel.

The *specificity* of a gene for a cell type is its HPA expression in that cell type divided by its summed expression across the 13 drawn cell types plus hepatocytes. Three filters were then applied. First, a gene had to reach ≥100 nCPM in the cell type it marks. Second, its expression in each stool-dominant cell type — neutrophils, colonocytes and goblet cells — had to be <5% of its expression in the marked cell type; a cell type was never judged against itself. Third, survivors were ranked by specificity in bins of 0.01, then by expression, and up to three were retained per cell type. All retained three except monocytes, which retained one (*VCAN*): monocyte and neutrophil transcriptomes overlap extensively (*CD14*, *FCN1*, *S100A12*, *SELL* and *CD163* are all high in neutrophils). Panels show log₁₀(TMM-CPM + 1) across the 77 Normal libraries beside each gene’s HPA expression across the major cell types of its own cluster and the stool-dominant cell types of the other, each row scaled to its maximum. Supplementary Fig. 6 gives the number of genes retained at each step.

### Lineage signature scores

The B-cell, T-cell (Fig. 4c) and neutrophil scores (Fig. 3b, Fig. 4a, d) are each the mean, over the genes of the set, of the gene’s z-score of log₂(TMM-CPM + 1), so that every gene contributes equally and the score is in standard-deviation units. All three sets are the curated panels specified a priori from canonical lineage biology: the B-cell set is the ten-gene B-cell panel (*MS4A1*, *CD79A*, *CD79B*, *CD19*, *PAX5*, *BANK1*, *BLK*, *CR2*, *TNFRSF13C* and *CD22*) and the T-cell set the 14-gene T-cell panel (*CD3D*, *CD3E*, *CD3G*, *CD2*, *TRAC*, *TRBC1*, *TRBC2*, *THEMIS*, *IL7R*, *LCK*, *ZAP70*, *CD28*, *IL17A* and *IL22*), both from the cross-tissue immune atlas^30^ and sorted-blood signatures^31^. The neutrophil set is the 15-gene panel above (*FCGR3B*, *CSF3R*, *CXCR1*, *CXCR2*, *FPR1*, *S100A8*, *S100A9*, *AQP9*, *TREM1*, *SELL*, *ITGAM*, *VNN2*, *CLEC4E*, *PROK2* and *TNFAIP6*: mature-neutrophil receptors, effectors and surface markers from sorted-population signatures^31^ and marker compendia^32^; *VNN2* encodes GPI-80, a surface protein of neutrophil secretory vesicles^35^).

### Antigen-receptor assembly

Antigen-receptor sequences were assembled with TRUST4 1.1.9^36^ from each library’s coordinate-sorted, duplicate-marked BAM file, using the TRUST4 hg38 BCR/TCR locus reference and the human IMGT^37^ V/D/J/C reference. Each clonotype was assigned to a locus from its V gene, or from its J or C gene where V was unassigned, and classed as productive if its CDR3 began with the conserved cysteine, ended with phenylalanine or tryptophan and contained no stop codon or frameshift. Of the 5,574 clonotypes in the 285 QC-passed libraries, 5,409 (97%) were productive.

### Antigen-database matching and invariant T-cell chains

CDR3 amino-acid sequences were reduced to their core (the IMGT junction without the conserved cysteine and phenylalanine or tryptophan) and matched exactly, within locus, against human records of VDJdb^38^ (release 2026-06-03), the IEDB^39^ TCR and BCR receptor tables (full export of 25 August 2026) and CoV-AbDab^40^ (downloaded 27 August 2026), 500,456 records in total. A CDR3 is counted once in each antigen class of its matching records, so class totals exceed the number of matched sequences.

Mucosal-associated invariant T (MAIT) cells carry a semi-invariant α chain joining *TRAV1-2* to *TRAJ33* or, less often, *TRAJ20* or *TRAJ12*^41,42^; in the TRUST4 IMGT reference these J segments encode DSNYQLI, SNDYKLS and MDSSYKLI before the anchor residue, each supplying the conserved Tyr95α that contacts MR1 and the ribityl moiety of the antigen^43^. MAIT-like α chains were defined as *TRAV1-2* joined to any of these J segments or, irrespective of V assignment, a CDR3α containing DSNYQLI; only *TRAJ33* was matched on sequence as well as on J assignment, because exonuclease trimming can shorten SNDYKLS to as little as YKLS. Chains with and without *TRAV1-2* are reported separately. Invariant natural killer T-cell α chains (*TRAV10*–*TRAJ18*) were sought the same way; none was found.

### Principal-component analysis

Log₂(TMM-CPM + 1) values of the 2,000 most variable genes were mean-centered per gene and decomposed by singular-value decomposition across the 271 libraries outside endoscopic remission (Normal 77, APL 63, CRC 55, IBD 76); eight components were retained. The sign of each component is arbitrary and was fixed as follows: PC1 was flipped where necessary so that it correlated positively (Spearman) with the neutrophil score; for every other component, the gene with the largest absolute loading was identified and the component was flipped where necessary so that this gene’s loading was positive. Fig. 3a, b place the 271 libraries on PC1 and the further component with the highest area under the receiver operating characteristic curve (AUC) for separating CRC from IBD, chosen by that rule (PC6 in this cohort), colored by diagnosis with group means marked by crosses and a 68% contour (1.51 s.d. ellipse) per group (Fig. 3a) or by the 15-gene neutrophil score, which is not an input to the decomposition (Fig. 3b; ρ, Spearman correlation of the score with each component over the 271 libraries). For Fig. 3c the scores of each of the eight components were standardized across all 271 libraries and their distributions shown for Normal, CRC and IBD, with the unadjusted Kruskal–Wallis P across the three groups; APL libraries enter the decomposition and the standardization but are not plotted. For each of the two plotted components, the 100 genes with the most positive and the 100 with the most negative loadings were tested for enrichment with Enrichr^44,45^ (via GSEApy^46^) against GO Biological Process 2023^47^, KEGG 2021 Human^48^, Reactome 2022^49^ and MSigDB Hallmark 2020^50^ (hypergeometric test, the 2,000 decomposed genes as background, Benjamini– Hochberg adjustment); the two most significant terms per pole are shown as −log₁₀ adjusted P against the adjusted P = 0.05 line, whether or not they reach it (Fig. 3d). Poles are labeled by component and sign; which diagnoses lie toward each pole is read from Fig. 3c.

### Differential expression

Differential expression between CRC and IBD used DESeq2^51^ (PyDESeq2^52^) on raw counts of the 55 CRC and 76 IBD libraries outside remission (131 in total), with the design ∼ condition, CRC as reference and Cook’s-distance outlier refitting. Before fitting, 448 nuclear-encoded mitochondrial pseudogenes and *MT-RNR2*-like genes were removed, as they distort size-factor estimation, and genes with fewer than 10 counts across the 131 libraries were discarded, leaving 27,315. Genes with Benjamini–Hochberg-adjusted P < 0.05 (Wald test) and |log₂ fold change| > 1 were called differentially expressed.

### Program scores

Four hallmark programs annotate the volcano plot of Fig. 3e and are scored in Fig. 3f, each declared in advance as one named term in the gene-set libraries above, on one side of the CRC-versus-IBD contrast: Wnt signaling (KEGG *Wnt signaling pathway*), epithelial– mesenchymal transition/stroma (Hallmark *Epithelial Mesenchymal Transition*) and proliferation/E2F (Hallmark *E2F Targets*) on the CRC side, and the interferon-γ response (Hallmark *Interferon Gamma Response*) on the IBD side. The genes differentially expressed on each side (522 higher in CRC, 769 higher in IBD) were tested with Enrichr against all tested genes as background, and each program is the overlap of its term with that list: 11, 23, 16 and 57 genes (Enrichr adjusted P = 0.019, 2.2 × 10⁻¹⁰, 1.9 × 10⁻⁵ and 3.0 × 10⁻³⁹). Because every member gene is selected by the contrast it annotates, the scores describe that contrast rather than provide independent evidence for it; they show whether the CRC–IBD difference is one axis or several programs. A program score is the mean z-score of log₂(TMM-CPM + 1) over its genes, standardized jointly across the Normal, IBD and CRC libraries (77, 76 and 55); CRC and IBD were compared by two-sided Mann–Whitney U test, with Normal shown for reference. Supplementary Fig. 8 shows each member gene by group.

### Classification

Classifiers were fitted on log₂(TMM-CPM + 1) by selecting the 2,000 most variable genes, standardizing, and fitting L2-penalized logistic regression (multinomial for more than two classes; lbfgs solver, C = 1.0, balanced class weights, up to 4,000 iterations), with gene selection and standardization refitted within each training fold. Cross-validation was five-fold, stratified by class and grouped by stool sample (scikit-learn^53^ StratifiedGroupKFold), and was repeated with ten random fold assignments; point estimates are means over the ten. Uncertainties are 95% percentile bootstrap intervals over libraries, resampled with replacement 2,000 times within class, the statistic on each resample being the mean over the ten fold assignments.

The three-class model was trained on CRC (55), IBD (76) and Normal (77) libraries (APL was excluded). Each library’s out-of-fold call is its consensus over the ten fold assignments, the class it was assigned most often, with ties broken by its mean out-of-fold probability; Fig. 3g is the confusion matrix of these calls, and its accuracy carries a 95% percentile bootstrap interval over libraries (2,000 resamples, class sizes kept). One-versus-rest AUCs are means over the ten fold assignments with the same bootstrap interval (Fig. 3h). CRC and IBD libraries whose consensus call was Normal were counted by tumor stage and endoscopic grade (Results). The 2,000-gene setting is where out-of-fold accuracy peaks over a sweep from 10 genes to all 58,320 (Supplementary Fig. 9; mean ± s.e. over the ten fold assignments).

A second model asked whether advanced neoplasia can be called from stool alone (Fig. 3i): CRC and APL pooled as the positive class (n = 118) against Normal (n = 77). Within each training fold the 20 genes with the largest absolute Welch *t* statistic between cases and Normal were selected, standardized and fitted by L2-penalized logistic regression (C = 1.0, unweighted classes); cross-validation was as above, with five fold assignments. The out-of-fold score, averaged over assignments, was read as CRC versus Normal and as APL versus Normal; each AUC carries a 95% bootstrap interval over libraries resampled within study group, and sensitivity at 90% specificity was computed within each assignment at the threshold leaving 90% of Normal libraries below it, then averaged. Genes selected in at least 20 of the 25 training folds are listed in the legend.

### IBD activity and fecal calprotectin

Activity analyses used the 66 graded IBD libraries of the activity-monitoring set (UC 33, CD 33), with the 24 healthy libraries as a reference band. Fig. 4a shows *S100A8* and *S100A9* as log₂(TMM-CPM + 1), the neutrophil score standardized within these 90 libraries, and the immune-cell share from the deconvolution (Fig. 1c, d) as a percentage of transcripts.

Associations with grade (remission < mild < moderate < severe) are Spearman correlations across the graded IBD libraries, healthy excluded, with UC and CD pooled in Fig. 4 and within subtype in the Results; for the neutrophil score, adjacent grades were also compared by unadjusted two-sided Mann–Whitney U test. Repertoire size (the number of productive clonotypes; Fig. 4b) and the B- and T-cell scores (Fig. 4c) were related to grade in the same way. Because the pooled grade combines the Mayo endoscopic subscore with SES-CD categories, instruments that score different lesions, each pooled correlation summarizes two diseases measured on different scales.

Fecal calprotectin (µg g⁻¹, routine clinical assay) was available for 32 UC and 31 CD libraries (cohort 1: 12 UC and 15 CD; cohort 2: 20 UC and 16 CD). Calprotectin and the neutrophil score were each correlated (Spearman) with grade on identical libraries, within subtype and cohort and with subtypes pooled (Fig. 4d; Supplementary Fig. 5), and with the raw endoscopic score (Supplementary Fig. 4), so that the two measures differ only in what is measured.

To ask whether activity can be called from the neutrophil score alone, a classifier was fitted on that single feature, standardized across the 90 libraries, to separate active disease (mild, moderate or severe; n = 54) from the quiescent gut, the 24 healthy and 12 remission libraries together (Fig. 4e). It uses the pipeline of *Classification* reduced to one feature: standardization, then L2-penalized logistic regression with balanced class weights, under five-fold cross-validation stratified by class and grouped by stool sample, with ten fold assignments. Fig. 4e shows its out-of-fold ROC as in Fig. 3h, with the AUC and its 95% bootstrap interval over libraries resampled within grade and the sensitivity at 90% specificity, as means over the ten fold assignments. With one feature the model is a monotone transform of the score, so the AUC is in effect the score’s own; the cross-validation places the call threshold (P ≥ 0.5), and hence its sensitivity and specificity, out of fold.

## Declarations

### Consent for Publication

All authors have approved the manuscript for submission. The content of this manuscript has not been published elsewhere.

### Data Availability

The raw sequencing datasets generated and analyzed during the current study are not publicly available due to patient confidentiality and the restrictions of the institutional ethical approval. However, de-identified data are available from the corresponding author upon reasonable request, subject to review and approval by the Guangdong Jiyin Biotech Data Access Committee.

### Author Contributions

Project conceptualization and study design: W.P., M.L. X.L., Z.Y, D.C..; Assay development and optimization: D.C., H.L. and J.H.; Development of the Python module for Bioinformatics and data analysis pipeline: W.P. and Y.L.; Clinical sample processing: D.C., H.L. and J.H.; Clinical data analysis: Y.L.; Patient recruitment, sample collection and patient data organization: Z.Y., X.L., M.L., S.H, C.S., B.L.; Project supervision: M.L and W.P.; Drafting and reviewing of the manuscript: all authors.

## Acknowledgements

The authors would like to thank the following people for their contributions: Bowei Han, Youjia Ma, and Yang Leng for their help with Bioinformatics pipeline; and Ruikun Huang, Bingkun Huang, Wei Tang, and Chaowei Fu for sample preprocessing; Xiaohua Zhang for sample coordination.

