## Supplemental file for "Stool host transcriptome reveals gut epithelial cell states and immune dynamics for gastrointestinal diagnosis and monitoring"

### Supplementary figures

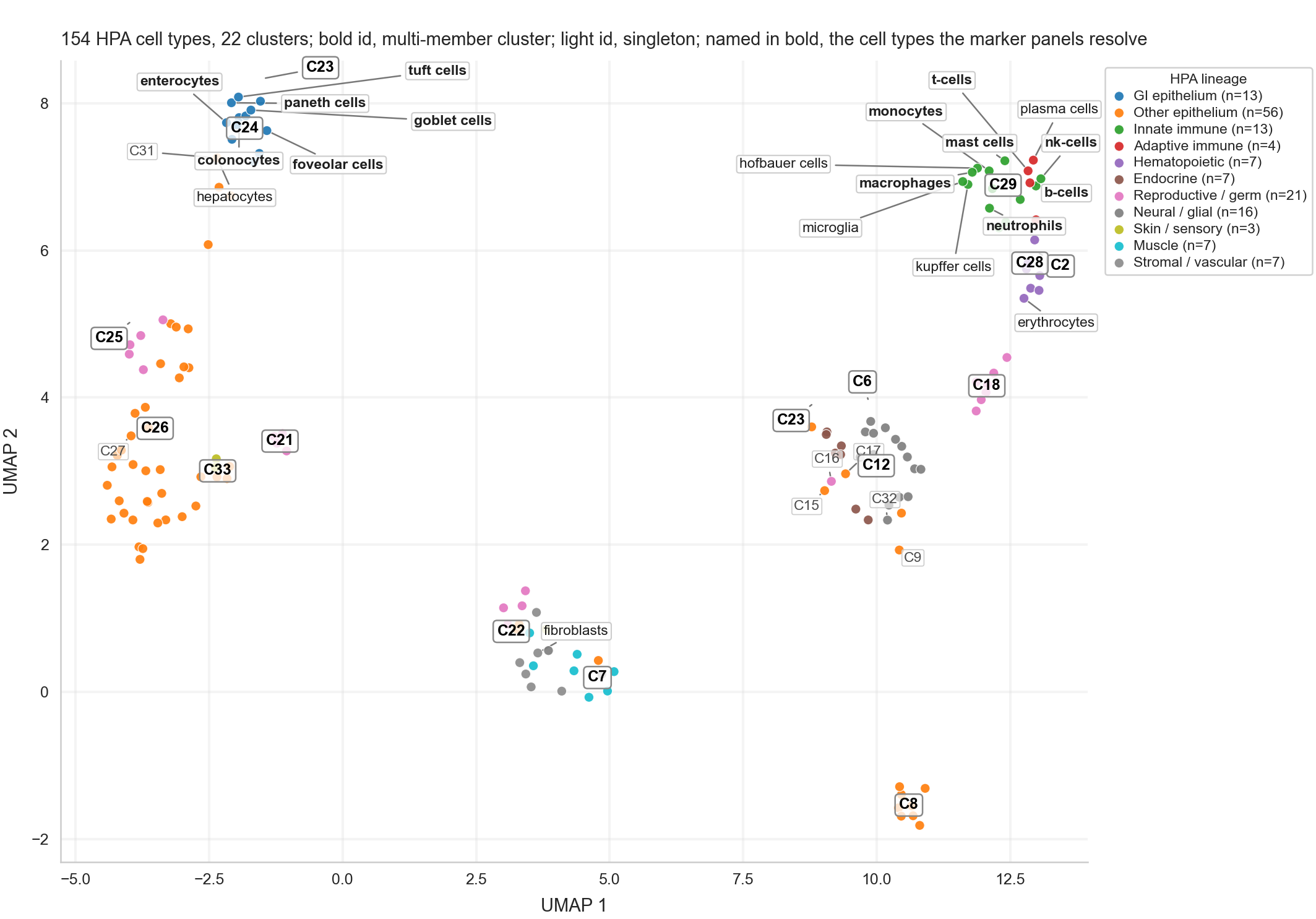

**Supplementary Figure 1 | The single-cell reference the deconvolution is built on.** UMAP of the 154 HPA single-cell types RNA-seq data on the 5,000 most variable genes (cosine metric, k = 10), colored by lineage. Bold for a cluster of several cell types, light for a singleton. The cell types named in bold are the thirteen the marker panels of Fig. 1e, f resolve; named in light type are hepatocytes and the tissue-resident myeloid types that sit beside the immune cluster's — Kupffer, Hofbauer and microglial cells — which is why monocytes and macrophages are hard to separate there. C24 is the columnar gastrointestinal epithelium, C31 beside it the hepatocyte singleton the fit carries as a liver column, and C29 the immune cells.

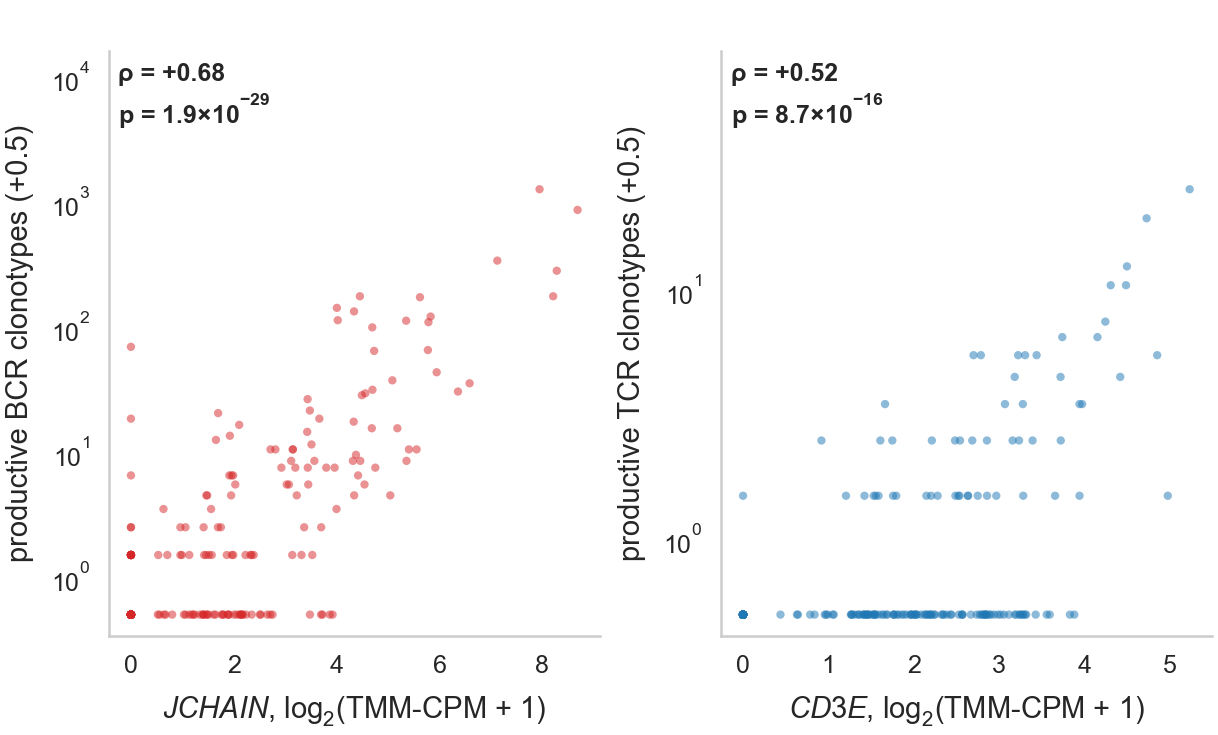

**Supplementary Figure 2 | Assembled antigen receptors repertoires are highly correlated with their lineage mRNA transcript.** Productive clonotypes per library against the matching lineage transcript across the 208 libraries of Fig. 2a, b (Normal, CRC and IBD outside endoscopic remission): BCR against *JCHAIN*, TCR against *CD3E* (Spearman).

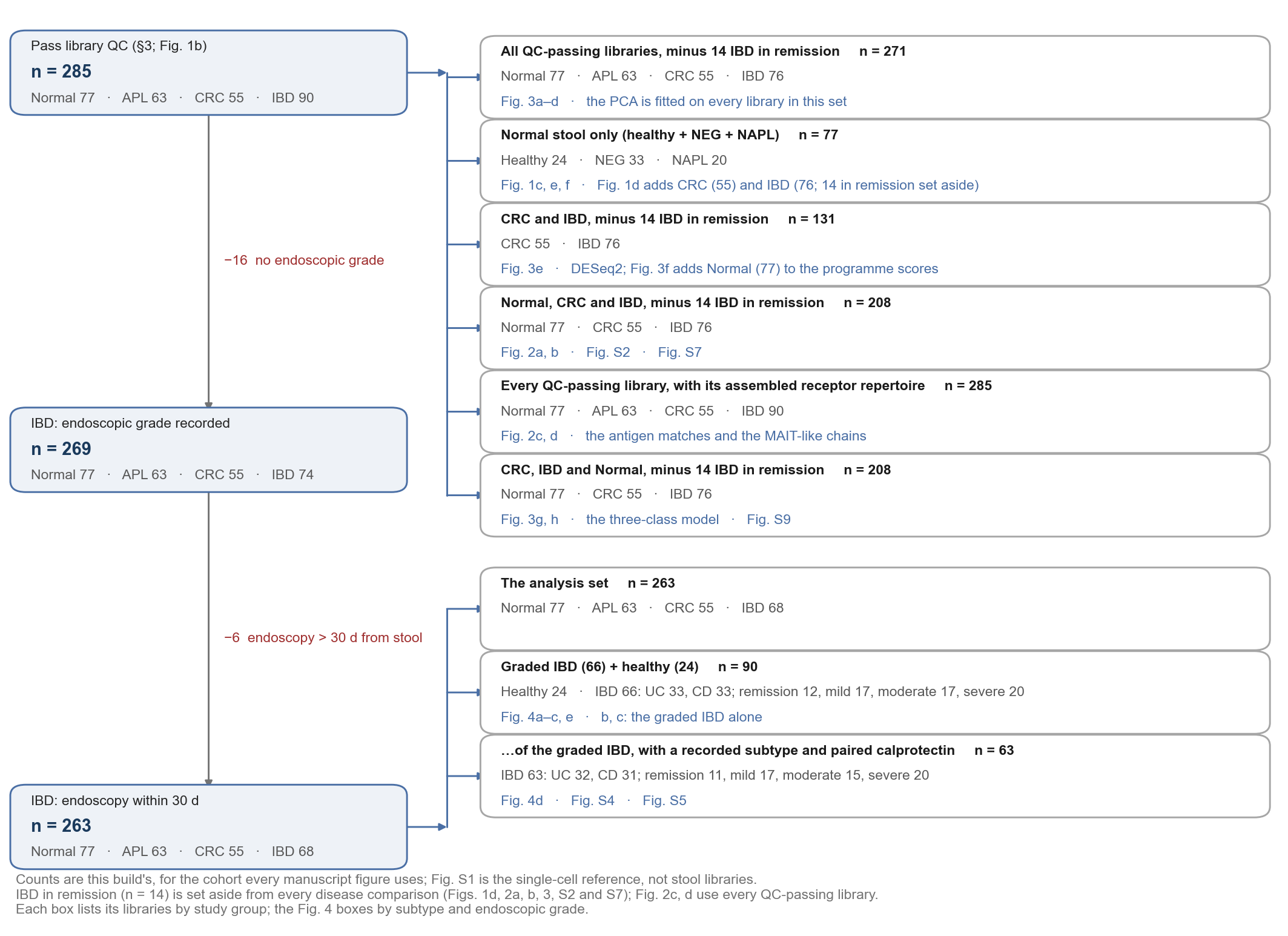

**Supplementary Figure 3 | The set of libraries behind each figure.** Left, the 285 libraries that passed library QC (Methods; Fig. 1b) and the two filters that define the activity-monitoring set, with the number removed at each step. Both filters apply only to IBD libraries, so a library of any other study group leaves the funnel at the first box and is unaffected by them. Right, the sets the figures are actually drawn on; each box lists its libraries by study group, and the Fig. 4 boxes by IBD subtype and endoscopic grade. The upper group is taken from the QC-passing libraries; Normal pools the healthy, colonoscopy-negative and non-advanced-lesion libraries in every set, and the principal-component analysis and the three-class model of Fig. 3 are fitted on every QC-passing library except the 14 IBD libraries in endoscopic remission, which are set aside from that figure only, so their N falls below the QC row of the funnel. The lower group is taken from the analysis set: Fig. 4a–c and e are drawn on its graded IBD libraries, **a** and **e** with the healthy reference band beside them, and Fig. 4d and Supplementary Figs. 4 and 5 on those of them that have a recorded subtype and a paired calprotectin.

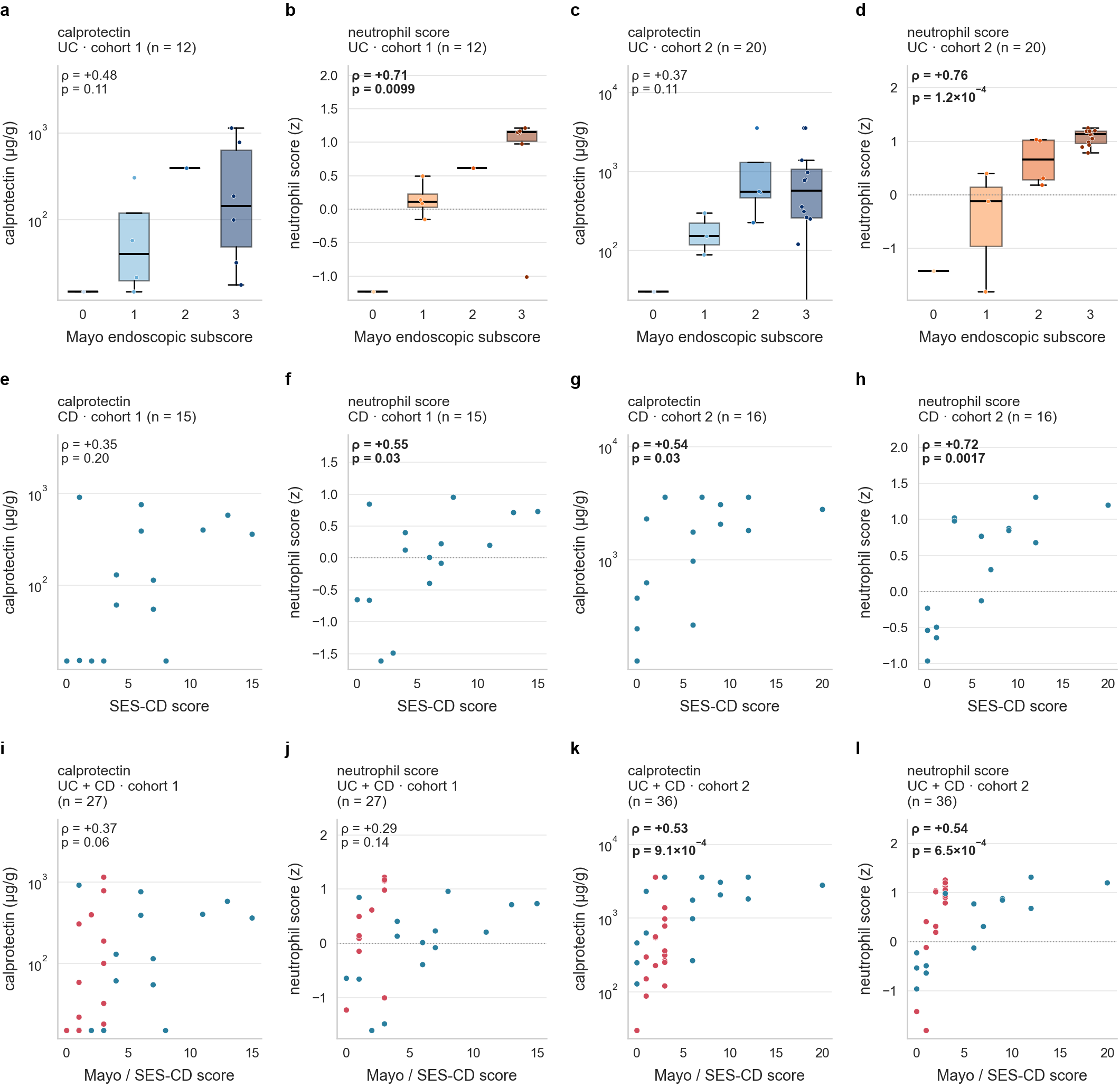

**Supplementary Figure 4 | Calprotectin and the stool neutrophil score against the raw endoscopic score, by subtype and recruiting cohort.** Clinical fecal calprotectin (**a**, **c**, **e**, **g**, **i**, **k**) and the 15-gene stool neutrophil score (**b**, **d**, **f**, **h**, **j**, **l**) against the endoscopic activity score, over the IBD libraries with a paired calprotectin and an endoscopic grade within 30 days of stool collection. Rows are ulcerative colitis (**a**–**d**), Crohn's disease (**e**–**h**) and the two pooled (**i**–**l**); within each row the first two panels are cohort 1 and the last two cohort 2. ρ is the Spearman rank correlation coefficient between the marker (calprotectin or the neutrophil score) and the endoscopic score, and p is the p-value of that correlation. Ulcerative colitis is scored by the Mayo endoscopic subscore (0–3), which has few enough values to draw as one box per score; Crohn's disease is scored by SES-CD, which runs 0–56 and spans 0–22 in this cohort, drawn as a scatter. **The pooled panels do not share one x-axis.** They place Mayo and SES-CD on the same scale although the two instruments score different organs and different lesions, so points are colored by subtype — red for ulcerative colitis, blue for Crohn's disease — and the pooled panels should not be read as a single severity scale. Supplementary Fig. 5 is the same subtype panels against the four-level grade band, and Fig. 4d the pooled row on that axis.

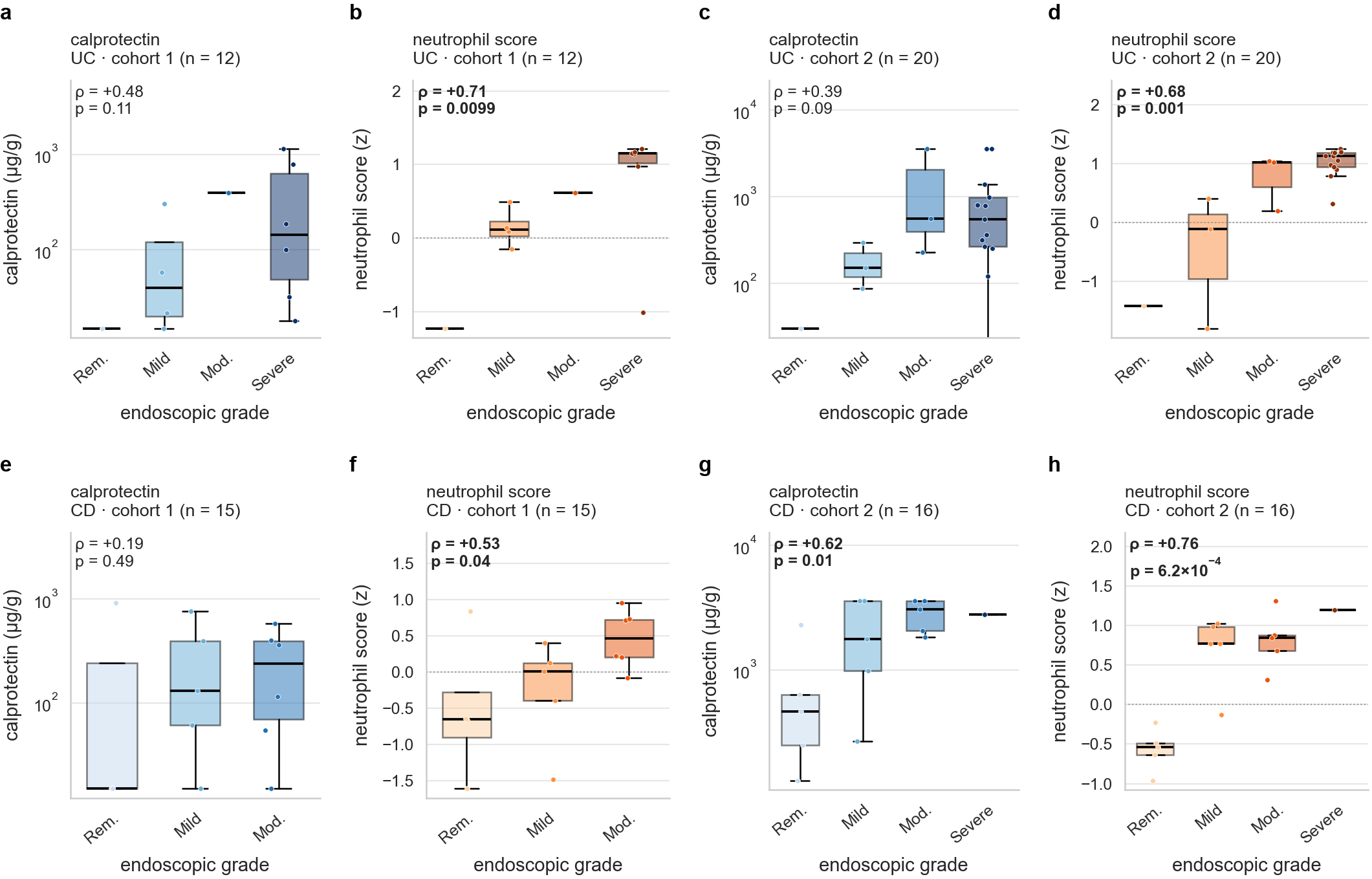

**Supplementary Figure 5 | Neutrophil score against endoscopic grade.** Panels, libraries, rows and columns are those of Supplementary Fig. 4 without its pooled row, which is Fig. 4d; only the x-axis differs, being the four-level endoscopic grade (remission, mild, moderate, severe) rather than the raw score. The stool neutrophil score outranks calprotectin in all six cohort × subtype cells: in ulcerative colitis ρ = +0.71 against +0.48 in cohort 1 and +0.68 against +0.39 in cohort 2; in Crohn's disease +0.53 against +0.19 in cohort 1 and +0.76 against +0.62 in cohort 2; pooled (Fig. 4d), +0.66 against +0.36 in cohort 1 and +0.75 against +0.11 in cohort 2.

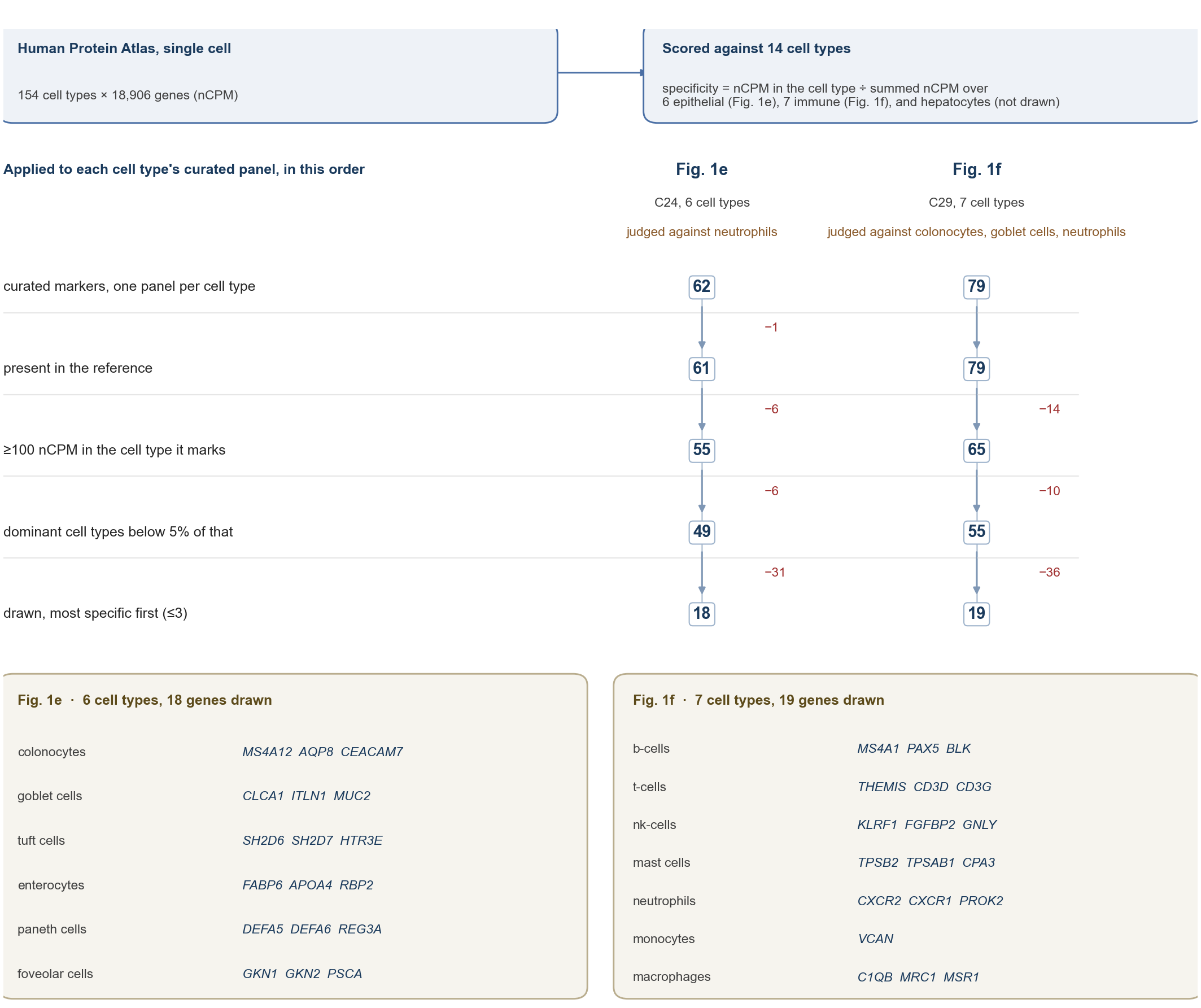

**Supplementary Figure 6 | How the marker genes drawn in Fig. 1e and 1f were chosen.** A flow chart of the selection, read from top to bottom. Top, the reference and the scoring rule that both panels share. Every gene is scored on the Human Protein Atlas single-cell reference against fourteen cell types: the six epithelial cell types of Fig. 1e, the seven immune cell types of Fig. 1f, and hepatocytes, which are scored against but not drawn. A gene's specificity for a cell type is its expression in that cell type divided by its summed expression over the fourteen. Middle, the filters, applied in order to each cell type's curated marker panel. The boxes give the number of genes left after each filter, separately for the epithelial panel (Fig. 1e) and the immune panel (Fig. 1f), and the red numbers give how many genes each filter removed. A gene must be present in the reference, reach 100 nCPM in the cell type it marks, and stay below 5% of that level in the cell types that dominate stool: neutrophils for the epithelial panel, and colonocytes, goblet cells and neutrophils for the immune panel, never counting the cell type being scored. The survivors are ranked by specificity and at most three are kept per cell type. Bottom, the genes drawn for each cell type. Monocytes keep one marker rather than three because monocyte and neutrophil transcriptomes overlap almost entirely and no threshold separates them; this is reported, not repaired. Every count is produced by the same function that makes the selection, so the chart is recomputed with each build and cannot disagree with the panels it explains.

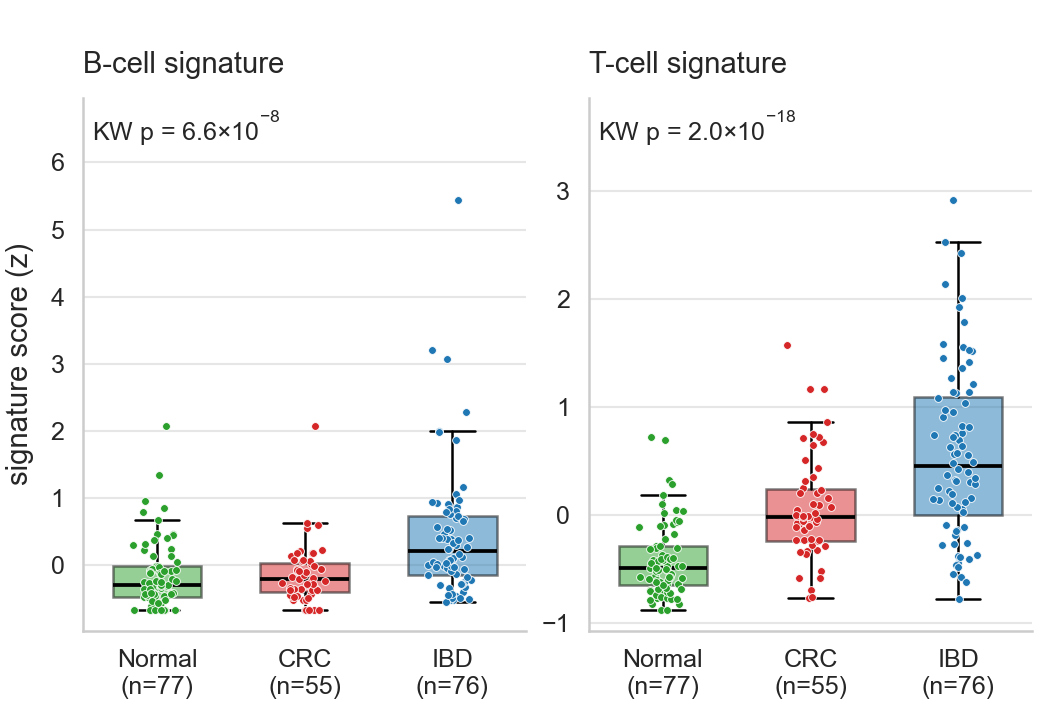

**Supplementary Figure 7 | B-cell and T-cell signature scores in Normal, CRC and IBD stool.** The B-cell signature (mean per-gene z-score of the ten-gene curated B-cell panel, *MS4A1*, *CD79A*, *CD79B*, *CD19*, *PAX5*, *BANK1*, *BLK*, *CR2*, *TNFRSF13C* and *CD22*, on log₂(TMM-CPM + 1) across the 285 QC-passed libraries) and the T-cell signature (the 14-gene curated T-cell panel, *CD3D*, *CD3E*, *CD3G*, *CD2*, *TRAC*, *TRBC1*, *TRBC2*, *THEMIS*, *IL7R*, *LCK*, *ZAP70*, *CD28*, *IL17A* and *IL22*), computed as in Methods, in the same Normal (n = 77), CRC (n = 55) and IBD (n = 76; endoscopic remission set aside) libraries as Fig. 2a, b. KW, Kruskal–Wallis over the three groups. The single transcripts *JCHAIN* and *CD3E* are Fig. 2b; the signatures reproduce their ordering — Normal below CRC below IBD — with tighter distributions. Boxes show median and interquartile range, whiskers 1.5 × IQR, points individual libraries.

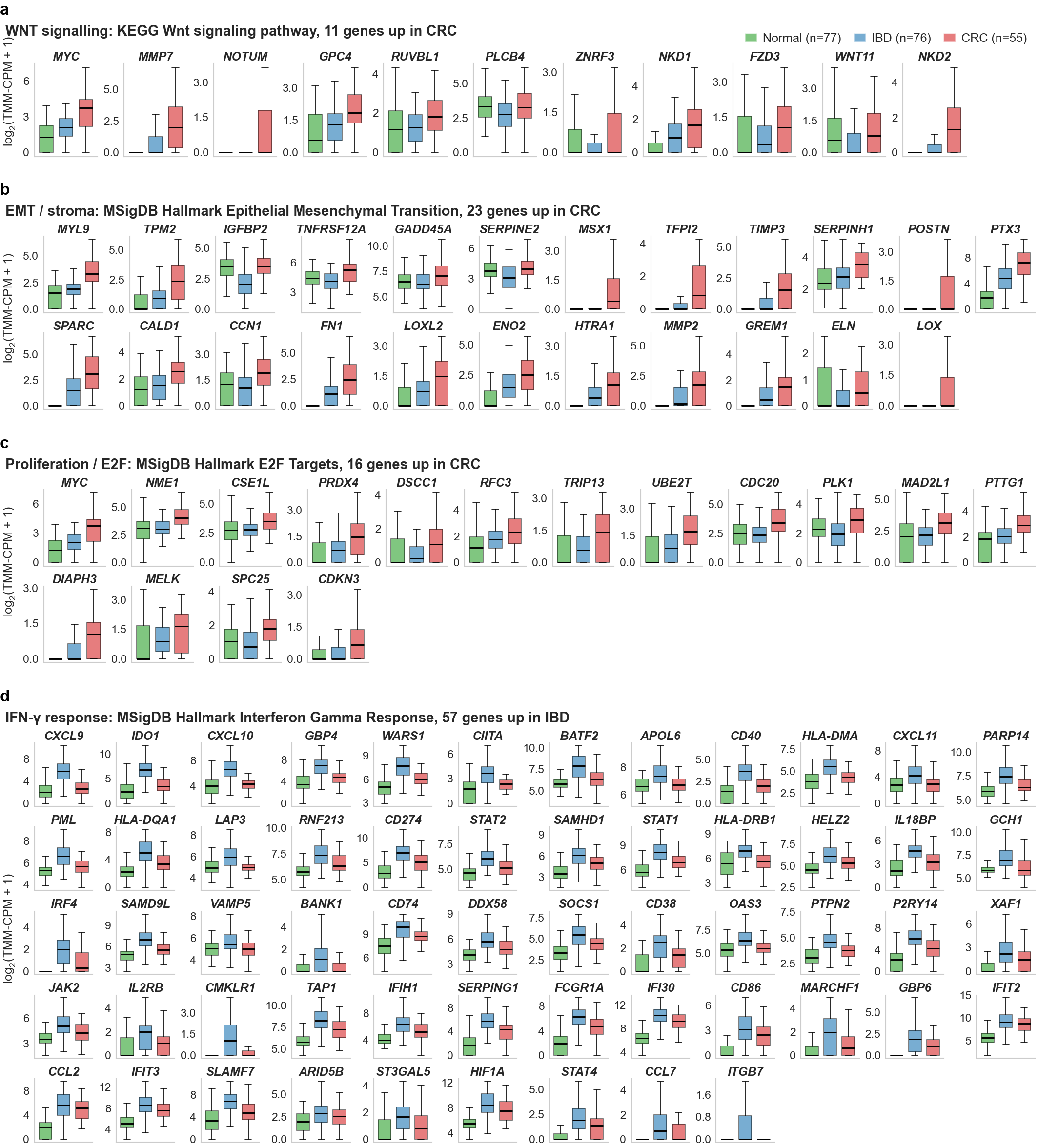

**Supplementary Figure 8 | Every gene behind the program scores of Fig. 3f.** One panel per program — **a**, WNT signaling (KEGG *Wnt signaling pathway*; 12 genes higher in CRC); **b**, epithelial–mesenchymal transition/stroma (MSigDB Hallmark *Epithelial Mesenchymal Transition*; 24 genes higher in CRC); **c**, proliferation/E2F (Hallmark *E2F Targets*; 15 genes higher in CRC); **d**, interferon-γ response (Hallmark *Interferon Gamma Response*; 58 genes higher in IBD) — each member gene as one box per group on log₂(TMM-CPM + 1), in the same Normal (n = 77), IBD (n = 80) and CRC (n = 55) libraries as Fig. 3f, genes in the order the CRC-versus-IBD contrast ranks them, most significant first. Every gene drawn is differentially expressed between CRC and IBD by construction (Methods); the panels add where Normal stool sits, which the contrast does not see, and whether a program score is carried by a few genes or by most. Boxes show median and interquartile range, whiskers 1.5 × IQR, without individual points.

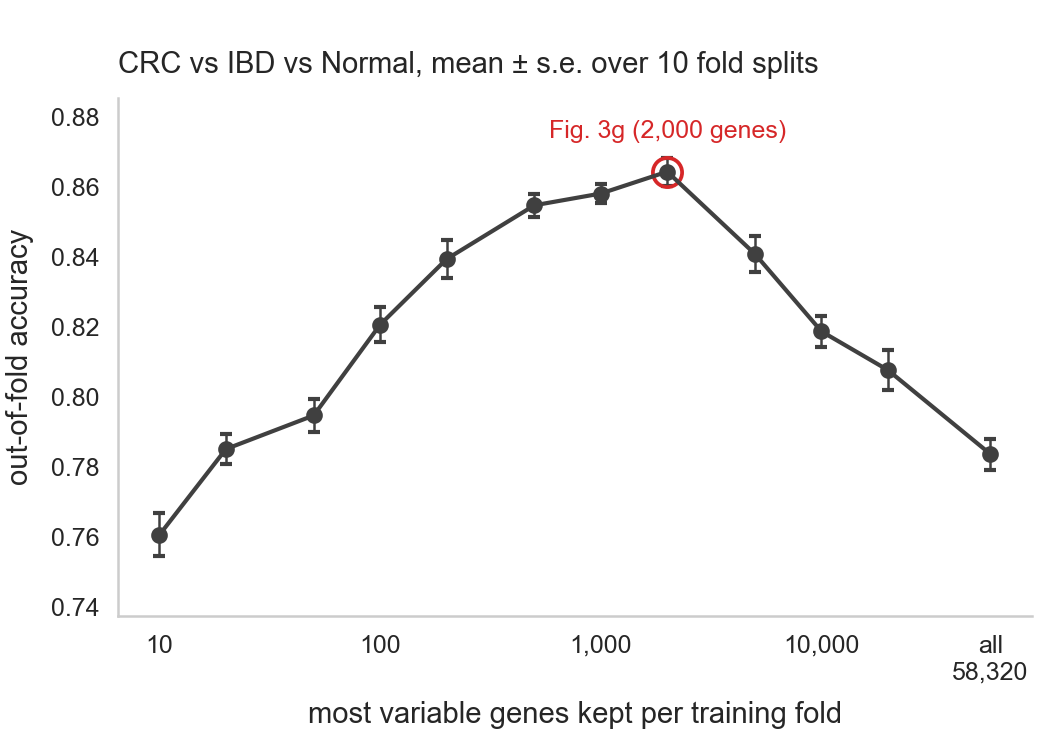

**Supplementary Figure 9 | Out-of-fold accuracy of the three-class model against the number of genes it uses as the features.** The CRC-versus-IBD-versus-Normal model of Fig. 3g refitted keeping 10 to all 58,320 genes per training fold (the most variable first, selected within the fold; classifier, folds and repeats otherwise as in Fig. 3g); the accuracy of each fold assignment's calls, mean ± s.e. over the ten assignments (Fig. 3g's title is instead the accuracy of the consensus calls). The circled point, 2,000 genes, is the model of Fig. 3g.

### Supplementary tables

**Supplementary Table 1 | The clusters of the single-cell reference behind the deconvolution of Fig. 1c, d.** One row per cluster of the Human Protein Atlas single-cell reference as the deconvolution used it (Methods, *Reference atlas and lineage clusters*): the cluster, its name (a trailing “+” marks a cluster whose members come from more than one lineage of the reference; a one-member cluster carries its cell type's name), the number and identity of the reference cell types it holds, whether it entered the fit, and the signature genes it was fitted on. A cluster's signature is the mean nCPM of its member cell types over these genes, chosen at cluster level by the three criteria of Methods (10 nCPM in the top cluster, 85% of the summed cluster means held by that cluster, expression in at least 34% of the cluster's members) with the cell-type-resolution check for the one-member clusters; the list is restricted to the genes present in the stool count matrix, which is what the fit sees. A cluster with fewer than five such genes is listed with the markers it had and marked as not fitted; the fitted clusters are the columns of the signature matrix, and their transcript shares are what Fig. 1c, d report. The table is written by the figure build from the same objects the fit uses.

| **cluster** | **cluster name** | **member cell types (n)** | **member cell types** | **fitted** | **signature genes (n)** | **signature genes** |
| --- | --- | --- | --- | --- | --- | --- |
| C02 | Hematopoietic | 2 | megakaryocytes; platelets | yes | 35 | PPBP; TREML1; LY6G6F; TUBB1; PF4; GP9; PF4V1; AQP10; PTCRA; NRGN; CMTM5; SCGB1C2; SCGB1C1; CLEC1B; OR2B6; RGS18; MPIG6B; ITGB3; GP5; NAT8B; TRAPPC3L; CAVIN2; SLFN14; GP1BA; GRAP2; SLC35D3; FERMT3; GNG8; TBXA2R; SELP; H3C10; PTGIR; ITGA2B; CDKN2D; BEND2 |
| C06 | Neural / glial | 2 | cone photoreceptor cells; rod photoreceptor cells | yes | 46 | OPN1LW; OPN1MW; RBP3; ARR3; RCVRN; PDE6H; AIPL1; PPEF2; MPP4; RHO; IMPG1; GRK7; TULP1; RS1; PDC; SMIM40; SAG; PDE6A; GUCA1A; RAB41; GNAT1; PDE6G; PRPH2; GNGT2; TMEM244; SAMD7; CPLX4; NR2E3; RTBDN; GUCA1B; RAX2; GUCY2F; ANKRD33; GUCA1C; RP1L1; IMPG2; CNGB1; KCNV2; PSKH2; CABP5; EYS; RPGRIP1; SLC17A7; RD3; CACNA1F; ROM1 |
| C07 | Muscle + | 6 | adipocytes; cardiomyocytes; epicardial cells; fibro-adipogenic progenitors; myonuclei; thymic myoid cells | yes | 35 | MYL1; MYH2; TECRL; C10orf71; MYH7; ASB10; SMYD1; XIRP1; NRAP; CSRP3; MYH1; TTN; MYH6; MYL2; FBXO40; NEB; KLHL38; TRDN; MYBPC3; MYBPC1; LMOD2; TCAP; SGCG; KLHL31; MYOT; STYXL2; RBM24; ANKRD2; MYOZ2; TRIM63; TNNI1; PPP1R3A; ACTA1; CASQ2; CKM |
| C08 | Other epithelium | 8 | distal convoluted tubule cells; loop of henle epithelial cells; papillary tip epithelial cells; podocytes; proximal tubule cells; renal collecting duct intercalated cells; renal collecting duct principal cells; renal connecting tubule cells | yes | 21 | SLC12A3; AQP2; UTP14C; UMOD; TRPV5; LHX1; MIOX; ZNF365; POU3F3; GCGR; PALM3; TIGAR; CNKSR3; KCNJ1; POU5F1; SOCS5; SLC14A2; UMAD1; HNF1B; SLC12A1; NAIP |
| C12 | Neural / glial + | 20 | astrocytes; bergmann glia; brain excitatory neurons; brain inhibitory neurons; corticotrophs; ependymal cells; gonadotrophs; lactotrophs; müller glia; oligodendrocyte progenitor cells; oligodendrocytes; other brain neurons; pituicytes/fscs; pituitary stem cells; retinal amacrine cells; retinal bipolar cells; retinal ganglion cells; retinal horizontal cells; somatotrophs; thyrotrophs | yes | 8 | PRL; PCDHA6; ZNF883; POMC; POU3F2; SLITRK1; KCNC2; DBNDD2 |
| C18 | Reproductive / germ | 7 | differentiating spermatogonia; early primary spermatocytes; early spermatids; late primary spermatocytes; late spermatids; oocytes; undifferentiated spermatogonia | yes | 50 | FAM47B; IQCF6; C20orf173; ZNRF4; PAGE3; PRKACG; MPC1L; ATXN3L; GAGE1; FAM9A; PRM2; ESX1; PRR30; C10orf120; VCX2; CCDC185; TNP1; PRM1; DUSP21; H2AP; FAM170A; ACTL7A; IQCF5; C20orf141; TEX47; FOXL3; SIRPD; HMGB4; MAGEA1; LELP1; CABS1; HSFX3; CT83; TEX13A; CYLC2; C16orf82; LYZL6; FAM187B; SSX3; BPIFA3; CCDC70; C16orf78; CCDC54; P3R3URF; CAPZA3; FTMT; CFAP97D1; ACTRT2; TEX38; H1-7 |
| C21 | Reproductive / germ | 4 | cytotrophoblasts; extravillous trophoblasts; migrating cytotrophoblasts; syncytiotrophoblasts | yes | 42 | CGB3; CSH1; INSL4; LGALS16; XAGE2; PAGE4; XAGE3; CGA; VGLL1; CSH2; PSG5; PSG2; GCM1; LAIR2; HSD3B1; DUSP9; PSG3; LGALS14; ISM2; LGALS13; ERVW-1; CYP19A1; CGB7; MAGEA8; PGF; SIGLEC6; ERVH48-1; SLC22A11; GJA5; EBI3; LEP; HTRA4; SLC52A1; PSG4; CAPN6; APELA; WNT7A; CALHM4; FSTL3; DLX4; HLA-G; KRT23 |
| C23 | GI epithelium + | 2 | neuroendocrine cells; pancreatic islet cells | yes | 28 | GCG; INS; MLN; INSL5; FEV; PYY; PPY; CRYBA2; SST; IAPP; KCNK16; UCN3; GHRL; GIP; NTS; CHGA; LCN15; TPH1; RXFP4; VGF; GPR119; GAST; SCT; REG4; FFAR1; CFC1; PAX4; NKX2-2 |
| C24 | Columnar GI epithelium | 12 | colonocytes; enteric stem cells; enteric transient amplifying cells; enterocytes; foveolar cells; gastric chief cells; gastric progenitor cells; goblet cells; mucous neck cells; paneth cells; parietal cells; tuft cells | yes | 19 | GKN1; ZG16; PGC; CCL25; GUCA2A; LIPF; FABP2; FABP6; GUCA2B; MUC17; FCGBP; TFF2; APOBEC1; TFF1; SPINK4; SI; RBP2; MYO1A; LGALS4 |
| C25 | Reproductive / germ | 2 | epididymal clear cells; epididymal principal cells | yes | 49 | DEFB113; DEFB128; DEFB114; DEFB118; PATE1; DEFB110; EDDM3B; DEFB129; SPAG11A; TEDDM1; DEFB112; DEFB127; RNASE9; WFDC9; SPAG11B; LCN6; LCN9; DEFB132; DEFB135; DEFB121; DEFB126; CLPSL2; EDDM3A; CST11; DEFB131A; PATE3; WFDC10A; LCN8; WFDC8; NPFF; DEFB108B; CD52; PATE2; ELSPBP1; BRS3; CITED1; CLPSL1; BSPH1; LCN12; CRISP1; PATE4; ADAM7; SPINK2; CAMP; SPINK13; LIPI; SPINT3; LRCOL1; ATP6V1G3 |
| C26 | Other epithelium + | 36 | alveolar cells type 1; alveolar cells type 2; basal prostatic cells; breast hormone-responsive cells; breast myoepithelial cells; breast secretory cells; cholangiocytes; conjunctival goblet cells; endometrial ciliated cells; endometrial glandular cells; endometrial luminal cells; endometrial secretory cells; epididymal basal cells; epididymal efferent duct absorptive cells; epididymal efferent duct ciliated cells; fallopian secretory cells; fallopian tube ciliated cells; lacrimal acinar cells; pancreatic acinar cells; pancreatic duct cells; prostatic club cells; prostatic glandular cells; prostatic hillock cells; respiratory basal cells; respiratory ciliated cells; respiratory deuterosomal cells; respiratory ionocytes; respiratory secretory cells; salivary acinar cells; salivary basal cells; salivary duct cells; salivary ionocytes; salivary myoepithelial cells; submucosal glandular cells; transitional alveolar cells; urothelial cells | yes | 5 | SCGB1A1; PIP; SCGB3A1; MSMB; BPIFB1 |
| C27 | breast lactating cells | 1 | breast lactating cells | yes | 6 | CSN2; LALBA; CSN1S1; CSN3; BTN1A1; CHRDL2 |
| C28 | Erythroid cells | 6 | erythrocyte progenitors; erythrocytes; hematopoietic stem cells; megakaryocyte progenitors; megakaryocyte-erythroid progenitors; thymocytes | yes | 9 | HBB; HBA2; HBA1; AHSP; SPTA1; KLF1; CRHBP; HBD; PRSS57 |
| C29 | Immune cells | 16 | b-cells; cdc; hofbauer cells; innate lymphoid cells; kupffer cells; macrophages; mast cells; microglia; monocyte progenitors; monocytes; neutrophil progenitors; neutrophils; nk-cells; pdcs; plasma cells; t-cells | yes | 35 | CCL3; RETN; FOLR2; CCL4L2; MARCO; CLEC4E; TLR8; C1QC; IL18RAP; FCGR3A; MMP9; P2RY13; MPEG1; GPR65; TLR7; BCL2A1; CCR2; TYROBP; ITGAX; MS4A6A; MS4A7; MNDA; OSM; NKG7; RNASE2; CYBB; C1QB; FCN1; FPR1; MS4A4A; CD86; HLA-DQA1; ITGB2; ALOX5AP; IGSF6 |
| C31 | hepatocytes | 1 | hepatocytes | yes | 41 | APOA5; MBL2; APOA2; CFHR2; CCL16; ORM1; F9; ITIH1; TAT; FGB; SPP2; SERPINC1; ORM2; SERPINA7; HPX; APCS; SAA4; F13B; FGA; HAMP; FGG; HRG; CFHR5; ALB; APOF; CFHR4; C8A; AGXT; C9; SLC25A47; SERPINA11; APOH; CYP2C9; HAO1; CYP2E1; AKR1C4; MAT1A; CYP2C8; BAAT; CPN2; CYP2A6 |
| C33 | Squamous epithelium | 6 | basal keratinocytes; esophageal apical cells; esophageal basal cells; esophageal suprabasal cells; ocular epithelial cells; suprabasal keratinocytes | yes | 50 | CRCT1; CRNN; SPRR1A; SPRR3; SLURP1; SPRR1B; SPRR2D; TMPRSS11B; SPRR2A; SBSN; SPINK7; KRT78; KRT4; SPRR2E; CYSRT1; DYNAP; KRT13; KRT6C; KRT6A; FAM25A; PGLYRP3; IGFL1; SPINK5; KRT6B; SPRR2B; GBP6; CNFN; LGALS7B; S100A7; IL36G; CSTA; KRT1; RHCG; PRSS27; KRT16; LGALS7; LY6D; IVL; DSG3; LYPD2; CALML3; S100A2; SCEL; KRT3; SERPINB13; ECM1; GJB2; DSG1; KLK12; GJB6 |
| C09 | choroid plexus epithelial cells | 1 | choroid plexus epithelial cells | no: 0 markers, fewer than 5 | 0 |  |
| C15 | adrenal cortex cells | 1 | adrenal cortex cells | no: 0 markers, fewer than 5 | 0 |  |
| C16 | sertoli cells | 1 | sertoli cells | no: 0 markers, fewer than 5 | 0 |  |
| C17 | adrenal medulla cells | 1 | adrenal medulla cells | no: 0 markers, fewer than 5 | 0 |  |
| C22 | Stromal / vascular + | 18 | decidual stromal cells; endometrial stromal cells; fibroblasts; granulosa cells; hepatic stellate cells; leydig cells; lymphatic endothelial cells; medullary thymic epithelial cells; melanocytes; mesothelial cells; myosatellite cells; ovarian stromal cells; pericytes; peritubular myoid cells; schwann cells; smooth muscle cells; vascular endothelial cells; vascular smooth muscle cells | no: 0 markers, fewer than 5 | 0 |  |
| C32 | retinal pigment epithelial cells | 1 | retinal pigment epithelial cells | no: 0 markers, fewer than 5 | 0 |  |
